# Incorporating Dietary Information to Enhance Polygenic Prediction Models with Applications to Body Mass Index and Type 2 Diabetes

**DOI:** 10.1101/2025.10.03.25337287

**Authors:** Eunice Y. Lee, Bryan L. Dinh, Ji Tang, Samantha Streicher, Xinran Wang, Subarna Biswas, He Tian, Xian Yu, Kekoa Tappara, Take Naseri, Satupa‘itea Viali, Daniel E. Weeks, Jenna C. Carlson, Christopher A. Haiman, Loic Le Marchand, Gertraud Maskarinec, Lynne R. Wilkens, Song-Yi Park, Charleston W.K. Chiang

## Abstract

Polygenic predictors can enhance screening for biomedical conditions, such as metabolism-related traits and diseases, but explain limited phenotypic variance and face implementation challenges in non-European populations. On the other hand, dietary quality and other sociocultural factors are well established metabolic risk factors that remain under-investigated in risk stratification models. In this study, we developed and evaluated risk stratification model combining polygenic predictors and diet-based models for body mass index (BMI) and type 2 diabetes (T2D). Using 5,368 Native Hawaiians from the Multiethnic Cohort (MEC-NH) with genetic data, we integrated large-scale cross-ancestry GWAS summary statistics to develop polygenic score (PGS) models with better prediction accuracies (partial-R^2^ [SE] = 0.12 [0.04] for BMI; liability-R^2^ [SE] = 0.09 [0.04] and AUC = 0.65 for T2D) than using GWAS information from single ancestry (partial-R^2^ = 0.03-0.09 for BMI and liability-R^2^ = 0.01-0.07 and AUC = 0.52-0.63 for T2D) or in combination with GWAS from MEC-NH (partial-R^2^ = 0.04-0.07 for BMI and liability-R^2^ = 0.01-0.04 and AUC = 0.53-0.62). Moreover, machine learning models trained on 520 dietary variables available from 14,344 MEC-NH individuals substantially explained BMI variation (partial-R^2^ [SE] = 0.12 [0.01]) and enhanced prediction when combined with PGS (adjusted-R^2^ = 0.29). The best performing diet score model for BMI was associated with multiple chronic diseases in the same cohort, potentially mediated via inflammatory and lipid pathways. Both PGS and dietary scores provided significant, complementary information for predicting BMI and T2D. For populations with limited genetic studies like Native Hawaiians, integrating external GWAS data or non-genetic dietary information can improve risk stratification models.

## INTRODUCTION

Metabolic syndrome (MetS), also known as insulin resistance syndrome, is defined by the presence of three or more co-occurring conditions: abdominal obesity, hypertension, hyperglycemia (1–2). These conditions increase the risk of developing cardiovascular disease (CVD), nephropathy, and type 2 diabetes (T2D), potentially leading to premature death (3–5). Currently, nearly 2.5 billion adults world-wide (43% of the world’s population) are overweight or obese (6). According to the CDC, obesity and T2D prevalence in both adults and children in the United States continues to increase and is projected to rise further (7–8). Early identification of risk factors can help prevent future incidence and reduce disease prevalence.

Genetic factors influence individuals’ responses to environmental stressors, causing them to react differently to the same health challenges. The within-family heritability estimates, which are relatively free of bias due to shared environment, are 0.43 for T2D and 0.49 for BMI across major ethnic groups in the United States (9). Hundreds to thousands of genetic variants discovered in recent large-scale genome-wide association studies of metabolic phenotypes across diverse populations demonstrate that these traits are polygenic. Rather than focusing on single genes or small gene sets, aggregating cumulative effects across the genome provides better risk prediction and stratification. Furthermore, polygenic scores (PGS) offer additional benefits, as adding PGS as covariates significantly increases disease prediction accuracy alongside other clinical factors (10–12). However, PGS face transferability issues across populations because their performance depends heavily on the underlying GWAS, which have been predominantly conducted in European-ancestry individuals (13–14). This results in low prediction accuracy for non-European populations, hindering clinical implementation and presenting an ongoing challenge (15–16).

Besides genetic contributions, MetS results from multiple environmental factors, with diet being one of the primary contributors. Research highlights that chronic low-grade inflammation, insulin resistance, and adipose tissue dysfunction are key pathogenic mechanisms in MetS. The intake of specific food components plays an important role in MetS development, prevention, and treatment (17–19). Unhealthy dietary habits such as consuming excess energy, sugar, saturated fats, and salt are major sources of low-density lipoprotein accumulation, promoting inflammation and altering glucose metabolism, thereby increasing diabetes risk and atherosclerotic plaque formation that can lead to cardiovascular conditions. A meta-analysis of 50 prospective and randomized controlled studies demonstrated that Mediterranean diet adherence reduced MetS incidence by 50%, indicating that healthy dietary patterns such as Mediterranean, DASH, vegetarian, or high-Healthy Eating Index diets can improve immune and cardiometabolic function while reducing chronic disease risk (20–22).

Similar to PGS, rather than examining individual food items or nutrients, dietary quality indices have been developed to assess overall dietary effects on chronic disease risk. However, these dietary indices are typically based on specific dietary recommendations and therefore do not capture the entirety of a person’s food choices and eating patterns, which are interactive and synergistic. Developing quantitative methods to detect eating behaviors and patterns that comprehensively reflect a person’s overall diet remains a major challenge. Moreover, these dietary indices usually tend to be developed to capture the overall health and well-being, rather than focusing on any specific disease or conditions. Therefore, individual dietary indices are likely to show weaker correlations with specific clinical outcomes.

In this study, we explored the role of genetic and non-genetic risk factors in MetS-related phenotypes including body mass index (BMI) and T2D by developing polygenic risk and diet scores in Native Hawaiians, a population with high prevalence of obesity and T2D (23–24). Moreover, there are limited available genetic data in this population, making polygenic predictions more challenging; non-genetic information such as dietary information is thus more pertinent in risk stratification in a clinical setting. For PGS development, we applied two recently developed Bayesian methods that integrate GWAS summary statistics from multiple populations, which have been reported to outperform existing PGS construction methods and improve cross-population polygenic prediction. For our diet score, we applied a data-driven approach to derive dietary patterns from all available food and nutrition consumption data using three different machine learning algorithms. Unlike traditional methods such as factor and cluster analysis, our data-driven approach considers both linear and non-linear relationships without making arbitrary decisions about factor or cluster structure. Our hypothesis-free method, requiring no prior knowledge, represents the totality of foods and drinks that individuals consume regularly and captures the synergistic effects of foods eaten together that may affect MetS outcomes. Combining both genetic and dietary information, our study has significant implications for disease prevention and precision medicine.

## METHODS

Overview of the methods, input data, and phenotypes for this study are described in Supplementary Figure S1.

### Study Subjects: Multiethnic Cohort

The Multiethnic Cohort (MEC) is a large-scale cohort designed to study diet and cancer among ethnic minorities in the United States. Study participants comprise 215,251 adult men and women aged 45 to 75 years at baseline who resided in Hawaii and California (primarily Los Angeles County). The cohort includes five racial and ethnic groups: Japanese Americans (28.5%), whites (24.8%), Latinos/Hispanic (22.7%), African Americans (16.8%), and Native Hawaiians (7.2%). Recruitment occurred from 1993 to 1996. Each participant completed a self-administered 26-page questionnaire that was mailed to them. A main portion of the questionnaire was a quantitative food frequency questionnaire (QFFQ) with more than 180 food items to assess habitual diet during the previous year (25). For each food item, the QFFQ queried consumption frequency using eight categories (“never or hardly ever”, “once/month”, “2-3 times/month”, “once/week”, “2-3 times/week”, “4 to 6 times/week”, “”once/day”, “≥2 times/day”) and portion size (three serving size choices). A calibration study demonstrated a good validity of the QFFQ against three 24-hour dietary recalls based on correlations in nutrient estimates (26). Genetic data were collected from a subset of volunteers who are slightly younger, less likely to smoke, and slightly more educated, but otherwise similar to the subset who did not donate biospecimen for genetics (**Table 1**). More information regarding the study design, questionnaire, and exclusion and inclusion criteria can be found elsewhere (25–27). The institutional review board of the University of Hawai‘I and University of Southern California approved the study protocol. All MEC participants signed an informed consent form.

**Table 1.** Descriptive statistics for Native Hawaiians in the Multiethnic Cohort (MEC). Never smoker, an adult who has never smoked or has smoked < 100 cigarettes in his or her lifetime; former smoker, an adult who has smoked ≥ 100 cigarettes in his or her lifetime but who had quit smoking at the time of interview; AC-METS, average of daily metabolic equivalent of physical activity during the last year (continuous); total energy intake, kcal/day. * Unpaired two-sample t-test and two-proportion z-test were conducted for continuous and categorical variables, respectively.

| Variable | Dietary data (n=14,344) | Genotype Data (n=5,374) | P-value* |
| --- | --- | --- | --- |
| **BMI (mean ± sd)** | 29.1 ± 6.3 | 28.9 ± 5.9 | 0.29 |
| **T2D Incidence (n,%)** | 4380 (30.5) | 1947 (36.2) | 2.74×10<sup>-14</sup> |
| **AGE (mean ± sd)** | 56.6 ± 8.7 | 55.0 ± 7.9 | <2.20×10<sup>-16</sup> |
| **SEX (n,%)** |  |  |  |
| Female | 8117 (56.6) | 3027 (56.3) | 0.75 |
| Male | 6227 (43.4) | 2347 (43.7) | 0.75 |
| **Smoking (n,%)** |  |  |  |
| Missing | 150 (1.0) | 52 (0.9) | 0.69 |
| Never Smoker | 5499 (38.3) | 2310 (43.0) | 3.10×10<sup>-9</sup> |
| Former Smoker | 5481 (38.2) | 2089 (38.9) | 0.40 |
| Current Smoker | 3214 (22.4) | 923 (17.2) | 1.11×10<sup>-15</sup> |
| **Physical Activity (mean ± sd)** |  |  |  |
| AC-METS | 1.7 ± 0.3 | 1.6 ± 0.3 | 0.04 |
| **Total energy intake (mean ± sd)** | 2601.9 ± 1342.6 | 2532.9 ± 1261.1 | 0.001 |
| **Education (n,%)** |  |  |  |
| Missing | 135 (0.9) | 53 (1.0) | 0.84 |
| < Middle School | 100 (0.7) | 14 (0.3) | 4.73×10<sup>-4</sup> |
| Middle School | 577 (4.0) | 101 (1.9) | 2.67×10<sup>-13</sup> |
| High School | 7023 (49.0) | 2206 (41.0) | <2.20×10<sup>-16</sup> |
| Vocational School | 1122 (7.8) | 459 (8.5) | 0.10 |
| Some College | 2991 (20.9) | 1326 (24.7) | 2.88×10<sup>-7</sup> |
| Graduated College | 1366 (9.5) | 702 (13.1) | 6.14×10<sup>-13</sup> |
| Graduate/professional School | 1030 (7.2) | 513 (9.5) | 4.33×10<sup>-8</sup> |
| **Ancestry (mean ± sd)** |  |  |  |
| EUR-like | NA | 0.28±0.26 | |
| EAS-like | NA | 0.26±0.26 | |
| *AFR-like* | *NA* | 0.01±0.03 |  |
| *PNS-like* |  | 0.45±0.23 |  |

The labels and groupings of the MEC individuals are based on self-report at baseline using terminology that was in practice in the early 1990s when the surveys were distributed to participants. In the survey, participants were asked to provide self-reported ethnic or racial background, marking all that applies, with the options of “Black or African-American”, “Chinese”, “Filipino”, “Hawaiian”, “Japanese (includes Okinawan)”, “Korean”, “Mexican or other Hispanic”, “White or Caucasian”, or “Other”. Most MEC analyses categorize participants into one of the five major racial/ethnic groups mentioned above that were targeted during recruitment, prioritizing group memberships in the following order: “African-American”, “Hawaiian”, “Latino”, “Japanese”, “White”. Thus, if an individual reports “Chinese” and “Hawaiian” in the survey, they would be classified as Native Hawaiian for these analyses. Among the 0.3% of individuals where no response was given for the participant, but existed for the mother and father, a race/ethnicity was created from the parent’s information; otherwise the individual is labeled as “Other.” While the MEC subcohorts represent multiple levels of self-reported ethnicities, nationality, and genetic ancestries, we continue to use these labels for consistency with other MEC studies and for ease of recognizing and interpreting the historical experience of health disparity for some of these populations, including potential benefits in genomic health through polygenic predictions. We recognize these labels are imperfect proxies for genetic ancestry, but note that our study focuses on a cohort of individuals from a community that emphasize self-identity or genealogical record for group membership (in place of estimated genetic ancestry), who have experienced significant disparity due to social determinants of health, who feel strongly of this identity, and who encompass multiple ancestries on a continuum that would be difficult and unethical to delineate or recategorize. As a convention we adopt in this study, we use the suffix *-like* when referring to genetic ancestry (*e.g.* Polynesian-like or PNS-like ancestry), and without the use of suffix when we are referring toa population of individuals (*e.g.* Native Hawaiians, Samoans, MEC-NH).

### Definition of Metabolic Health Phenotypes

#### Obesity/BMI

BMI was calculated using height and weight reported on the questionnaire at the entry of cohort for each participant. The raw BMI was first stratified by sex and regressed against the age. The residuals of the sex stratified regression models were inverse normalized and combined across sexes for the analyses. This residualized and inverse-normalized BMI is used throughout the study, and for brevity we simply refer to it as BMI.

#### Type 2 Diabetes

T2D status was ascertained from a few major sources of information for the MEC. First, self-reports were available from 6 questionnaires, in response to the question “Has your doctor ever told you that you had diabetes?” Second, as part of the biorepository, cohort members who reported T2D medication at time of blood draw were classified as cases. Third, a one-time linkage to Hawai‘i insurance databases as of Jan. 1, 2008, also indicated T2D cases for participants recruited in Hawai‘i. Finally, a linkage to Medicare Chronic Condition Warehouse for diabetes for Medicare claims between the years 1999-2019 (the latest linkage). Individuals indicated on any of these sources are considered a case, with the remaining individuals considered as controls. Our only information of prevalent diabetes is based on self-reports on the baseline questionnaire, and thus cases who did not report diabetes in the baseline questionnaire is considered incident T2D. For Native Hawaiians, we identified 6,566 total individuals with T2D (2,561 with genetic and dietary data, 4,005 with dietary data only). Of these, 4,380 are defined as incident T2D (1,947 with genetic and dietary data, 2,799 with dietary data only). 8,984 are defined as controls (2,433 with genetic and dietary data, 4,979 with dietary data only).

### Genotype Data Generation, Processing, and Quality Control

In total, 5,388 self-identified Native Hawaiians from the MEC were genotyped on two separate GWAS arrays: Illumina MEGA (n = 4,148) and Illumina Global Diversity Array (GDA; n = 1,226). Each dataset was processed through the same quality control pipeline: we removed variants that were genotyped in fewer than 95% of people in the sample, as well as variants out of Hardy-Weinberg equilibrium (p < 10^−6^). We also applied a filter for people with more than 2% missing genotypes but removed no one with this filter. The datasets were then lifted to hg38 using triple-LiftOver (v.1.33) (28) to ensure that alleles in inverted sequences between reference genome builds were properly lifted. We phased the genotypes with EAGLE (v.2.4.1) (29) using a Native-Hawaiian specific genetic map (30), and then imputed using the TOPMed imputation server against the TOPMed reference panel (release 3) (31).

Global ancestry estimates for the MEC-NH cohort were previously calculated (15) using ADMIXTURE (v.1.3.0) in unsupervised mode, in combination with other MEC ethnic groups and the 1000 Genome Project reference populations. We found MEC-NH individuals at k = 4 exhibited known components of European (EUR)-like, East-Asian (EAS)-like, and African (AFR)-like ancestries (interpreted based on 1000 Genomes reference samples), as well as a component of ancestry that is unique to the MEC-NH, presumed to be Polynesian (PNS)-like in origin.

We performed GWAS for BMI and T2D (prevalent + incident T2D cases) based on the subset of 4,148 individuals genotyped on the MEGA array, leaving out the 1,226 individuals genotyped on the GDA array as validation dataset. Specifically, we implemented the UW-GAC TOPMed GENESIS analysis pipeline (https://github.com/UW-GAC/analysis_pipeline) and inferred a Genetic Relationship Matrix (GRM) and Principal Components (PCs) using array data for MEC-NH. In brief, this pipeline filtered variants (MAF < 1%, missingness > 1%) before LD pruning (R^2^ > 0.1, 10Mb window). Initial kinship and population divergence estimates are inferred with KING (32). These estimates were used as input for PC-AiR to infer PCs that in turn served as input into PC-Relate to infer a genetic-relatedness matrix (33–34). A second iteration of PC-AiR and PC-Relate was run with the PC-Relate GRM as the input. Association testing is done with the GENESIS package (35). The null model is fit using the phenotype, GRM, and 20 PCs. Association testing is done with imputed dosages.

The Samoan dataset was downloaded from dbGAP (accession number: phs000914.v1.p1), containing 3,501 individuals genotyped at over 900,000 SNPs on the Affymetrix 6.0 array. The same quality control, phasing, and imputation procedures were followed.

### Trans-ethnic Polygenic Score (PGS)

The PGS were developed for each phenotype in MEC-NH participants applying two Bayesian approaches, namely PRS-CSx and BridgePRS, which are described below (36–37). For the development of BMI PGS, GWAS summary statistics conducted in individuals from the Taiwan Biobank and Biobank Japan (n=256,450) representing EAS-like ancestry, Genetic Investigation of Anthropometric Traits (GIANT) consortium and U.K. BioBank (n=∼700,000) representing EUR-like ancestry, and Native Hawaiians in MEC study (n=4,148 genotyped on MEGA array only) representing largely PNS-like ancestry (**Supplementary Figure S2)** were used. For the development of T2D PGS, we used ancestry-specific GWAS summary statistics from EAS-like populations (n=427,504) and EUR-like ancestry populations (n=1,812,017) as released by prior trans-ancestry GWAS efforts for T2D (38). We conducted GWAS for T2D in 4,131 MEC-NH in-house.

Pairs of these GWAS summary statistics were integrated and trained in both PRS-CSx and BridgePRS models separately. PGS were calculated based on the resulting model for validation in held-out MEC-NH (those genotyped on GDA array) for each phenotype (n_BMI_ = 1226; n_T2D_ = 1218); scores were standardized to have mean = 0 and standard deviation = 1. Following previous study (15), we measured performance of a PGS model based on the partial R^2^ (for continuous outcome, calculated by R package rsq v2.5) or the Nagelkerke pseudo-R^2^ (calculated by R package DescTools v0.99.43) transformed to the liability scale assuming an estimated lifetime risk of 60% (16, 39). Depending on the relative performance between PRS-CSx and BridgePRS models, the better-performing PGS by partial R^2^ was then used in the downstream analyses for the respective phenotype.

#### PRScsx

PRS-CSx utilizes a high-dimensional Bayesian regression framework to perform cross-population polygenic prediction by integrating GWAS summary statistics from multiple populations (36). For effect size estimation, information is shared between summary statistics while considering genetic diversity across discovery samples through population-specific allele frequencies and LD patterns. PRS-CSx applies a shared continuous shrinkage prior, which allows varying degrees of shrinkage to be applied to SNP effect sizes based on the strength of their GWAS associations. First, summary statistics and ancestry-matched LD reference panels were used to estimate population-specific posterior SNP effect sizes using PRS-CSx in auto mode, with default Strawderman-Berger prior parameters (a=1, b=0.5) for local shrinkage and the global shrinkage parameter (ɸ) learned from data using a fully Bayesian approach. The population-specific posterior effect estimates were meta-analyzed using inverse-variance weighting within the Gibbs sampler to produce combined SNP effects. The polygenic score for each study subject was generated using PLINK commands (--score and sum modifier that combines chromosome-specific scores into genome-wide scores).

We used pre-compiled 1000 Genomes reference panels provided by PRS-CSx for EAS-like and EUR-like ancestry summary statistics (∼1.2 million SNPs across multiple populations), and constructed a Native Hawaiian reference panel that conforms to the pre-compiled reference panel format. For the Native Hawaiian reference panel construction, we extracted the imputed genotypes at HapMap3 variants from a randomly chosen subset of 500 Native Hawaiian samples, restricted to variants with imputation quality filtering (R² > 0.8), MAF filtering (>1%), and back-converted the genome build to hg19 to be consistent with the pre-compiled 1000 Genomes references. We computed LD correlation matrices using PLINK and organized them into LD blocks using Asian population boundaries derived from population-specific LD breakpoints estimated by LDetect (40). The final reference panel includes population-specific allele frequencies and strand-flipping indicators integrated into the standard PRS-CSx multi-population framework, with LD matrices stored in compressed HDF5 format for computational efficiency.

#### BridgePRS

While PRS-CSx constructs PGS using continuous shrinkage priors applied at individual causal SNPs, BridgePRS operates at the locus level, combining SNP effects within genomic loci through Bayesian posterior mean estimation rather than simple averaging (37). SNP effects within each locus are modeled by a multivariate Gaussian distribution, where a locus is defined as variants with R² > 0.01 and within 1Mb of a lead variant (the SNP with the lowest p-value in that genomic region). We constructed three different trans-ancestry models for each phenotype: 1) EAS-like population as discovery and Native Hawaiian as target, 2) EUR-like population as discovery and Native Hawaiian as target, and 3) EAS-like population as discovery and EUR-like population as target. Single-ancestry or population-specific PRS models were trained and optimized using default parameters (α = 0.5 for allele frequency dependency, λ⁽⁰⁾ = 0.5 for baseline shrinkage), with zero-centered Gaussian prior distributions assigned to SNP effects at each locus. In the two-stage cross-ancestry models, posterior SNP effect sizes estimated in the discovery populations were used as informative priors for the target population (τ = 100 for cross-ancestry shrinkage). The final PRS combined all single-ancestry and two-stage polygenic scores using optimal weights determined through cross-validation ridge regression in the validation dataset (n_BMI=1,226; n_T2D=1,218). For EAS-like and EUR-like populations, we used LD information based on the EAS and EUR populations from 1000 Genomes, respectively, using variants with MAF > 5%. For Native Hawaiians, we computed the LD information based on 500 randomly selected individuals from the MEC-NH (the same set of individuals used above to construct the reference panel for PRS-CSx models).

### Dietary Variables and Phenotype Specific Diet Scores

Based on the QFFQ responses, participants’ food and nutrient intakes were computed using comprehensive food composition database developed and maintained for the MEC, including a large recipe database (25). As part of the Dietary Patterns Methods Project that the MEC and two other large US cohort studies participated, four commonly used indices (HEI, AHEI, aMED, DASH) for overall diet quality were calculated using a standardized methodology across the three cohorts (41). Later, 27 more dietary indices were additionally calculated. MEC dietary data includes nutrients (absolute intake), individual foods (grams), food groups (grams), MyPyramid Equivalents Database (MPED), and dietary patterns (component scores and total scores), which resulted in a total of 520 diet variables that were used to train and predict diet scores in MEC-NH subjects (n=13,595). For the phenotype-specific diet score, we applied an elastic net regularization, random forest, and neural network machine learning algorithms to prioritize diet variables or build regression models with dietary variables, from which we can compute a diet score that was then standardized within cohort to have a mean of 0 and standard deviation of 1. Across all diet score prediction models in Native Hawaiians, subjects without genetics data served as the training set and subjects with genetics data served as the validation set.

For the development and selection of the best-performing diet score, we applied three different machine learning algorithms. First, an elastic net regularization was implemented with a two-step nested cross-validation approach (5-fold cross validation) that identified the optimal L2 penalty parameter by evaluating cross-validation error across the lambda2 grid which ranged from 0.00001 to 0.01 with increments of 0.001, then the corresponding optimal L1 parameter for the selected L2 value was determined. After training the elastic net model on BMI (n=8,372), a single composite score was calculated for each participant in the validation dataset (n=5,223) based on the learned coefficients of the selected dietary variables. For the prediction of the diabetes outcome, a cross-validated elastic net logistic regression model was developed using the optimal lambda value identified through cross-validation, keeping the alpha fixed at 0.3 (blend of L1 and L2 penalties: 30% Lasso penalty for variable selection and 70% Ridge penalty for handling correlated predictors) in the training dataset (n= 7,026; 2,294 cases, 4,732 controls). Individual dietary scores were computed based on the elastic net model coefficients for the validation dataset (n= 4,632). We used the ’gcdnet’ (v.1.0.6) in R for the elastic net models

Second, a random forest regression model was trained to predict BMI using all dietary variables in the training dataset, configured with 500 decision trees. The trained random forest model was then applied to predict BMI values for each subject in the validation dataset. For diabetes status, a random forest classification model was trained to predict incident T2D status using all dietary variables in the training dataset. We applied the trained random forest model to the validation dataset to predict the probability of each individual having diabetes rather than providing binary classifications, and used the predicted probabilities as diabetes risk scores. For the random forest models, we used randomForest (v.4.7-1.2) in R.

Third, we applied a neural network diet score model. Multilayer perceptrons (MLPs) possess sufficient representational capacity to model the complex nonlinear relationships in terms of the Universal Approximation Theorem (42), and it is a common practice to use MLPs to fit a classifier and regressor in the machine learning field. The predictions of T2D and BMI based on dietary variables are technically a classification task and a regression-fitting task, respectively. Therefore, we employed MLPs to build neural network models for predicting T2D and BMI. Since the training is fast (one epoch takes seconds), we used grid-search to find the optimal values of the hyperparameters. Specifically, we defined multiple candidate values for the hyperparameters (**Supplementary Table S1**; the combination of these candidate values results in more than 1,500 candidate models for predicting BMI and T2D, respectively). We trained all the candidate models in parallel and chose the ones with the lowest validation loss as the final models. The key details for the training include: (a) 80% of the training data was used to optimize the weights and bias in the neurons, and the rest 20% of the training data was used as validation set to evaluate the validation loss; (b) The initial Learning rate was 0.01, and was multiplied by 0.5 if validation loss was not improved for 10 epochs; (c) Early stop was used to stop training after no improvement in validation loss for 15 epochs; (d) At most 200 epochs of training were executed; (e) Batch Normalization was used to normalize the outputs of each hidden layer; (f) Mini-batch training was used; (g) Dropout regularization with a rate of 0.1 was applied to hidden layers. As (c), (e), and (g) work together to effectively avoid overfitting, making it unnecessary to use the L1 and L2 regularizations further. The Keras within Tensorflow (43) was used to implement the building and training of the models.

### Mediation Analysis (Biomarkers)

We tested the association of the best-performing BMI diet score with the incidence of different chronic diseases ascertained from linkage of ICD codes from Medicare Claims with the Chronic Condition Warehouse in MEC-NH (44), adjusting for age, sex, BMI, total energy intake (kcal/day), and the first 10 genetic PCs. These chronic conditions included Alzheimer’s disease, anemia, asthma, atrial fibrillation, cataracts, chronic kidney disease, chronic obstructive pulmonary disease (COPD), depression, gout, hypertension and related phenotypes, ischemic heart disease, osteoporosis, rheumatoid arthritis, and stroke. For disease outcomes that were significantly associated with diet score, we conducted causal mediation analysis to examine whether levels of four different biomarkers (C-reactive protein (CRP), low-density and high-density lipoproteins (LDL and HDL), triglyceride) mediate the relationship between diet score and disease status. We employed a two-stage regression approach within the counterfactual framework for causal mediation analysis using the mediation R package (version 4.5.1). In the first stage, we fitted a linear regression model to estimate the mediator model, regressing in turn each biomarker on diet score while controlling for BMI, daily energy intake, and the first 10 PCs. In the second stage, we fitted a probit regression model for the binary disease outcome, regressing disease phenotype on both the biomarker (mediator) and diet score (treatment), while adjusting for age, sex, BMI, daily energy intake, and the same 10 PCs. We used nonparametric bootstrap resampling with 100 simulations to generate confidence intervals for the mediation effects. The analysis estimated the Average Causal Mediation Effect (ACME), Average Direct Effect (ADE), and total effect, representing the indirect effect through the biomarker, the direct effect of diet score, and the overall effect, respectively. Biomarkers measured in a subset of MEC-NH individuals included C-reactive protein (n=1,929), low-density lipoprotein (n=1,886), high-density lipoprotein (n=1,898), and triglycerides (n=1,894). Phenotype transformation for these biomarkers were performed as previously described (45).

## RESULTS

Baseline characteristics for the Native Hawaiian participants are shown in **Table 1**. The total number of subjects with dietary data was 14,344 and a subset of these individuals had their DNA samples genotyped (n=5,374). There were more female participants than males. Subjects with genetics data were slightly younger and had higher education levels than the overall group. All study subjects had their BMI information. The incidence of T2D from the enrollment to the latest follow-up was 4,380 and the subject with genetics data had significantly higher rate of T2D (36.2%).

### Developing the trans-ancestry PGS for BMI and T2D

To improve upon single-population PGS models previously developed for Native Hawaiians (15), we used two approaches (PRS-CSx and BridgePRS) to develop trans-ancestry PGS for BMI and T2D based on pairwise combinations of GWAS summary statistics from three continental populations: East Asian (n= 256,450 for BMI, 427,504 for T2D), European (n = 700,000 for BMI, 1,812,017 for T2D), and Native Hawaiians (n = 4,148 for BMI, 4,131 for T2D) **(Methods**).

For BMI, the best-performing trans-ancestry model with the highest accuracy in held-out validation MEC-NH (n = 1,226) individuals is one derived by PRS-CSx, combining GWAS summary statistics from EUR-like and EAS-like populations (927,557 common SNPs included in the model, with partial R^2^ = 0.119; **Figure 1**; Supplementary Table S2). This model outperformed single-population models previously constructed using pruning-and-threshold or LDPred2 (15), publicly available models (15), as well as models estimated by PRS-CS here (partial R^2^ = 0.029-0.093; **Figure 1**; Supplementary Table S2). Notably, in single-population PGS models using Native Hawaiian GWAS did not perform better than single-population PGS using East-Asian or European GWAS (partial R^2^ = 0.029 vs. 0.074-0.093; **Figure 1**; Supplementary Table S2), presumably due to its small size. In trans-ancestry PGS models, using Native Hawaiian GWAS in combination with East-Asian or European GWAS appeared to introduce more noise and lowered the accuracy of PGS models, compared to single-population PGS using either GWAS (partial R^2^ = 0.036-0.065 vs. 0.074-0.093; **Figure 1**; Supplementary Table S2). In fact, when we compared the two-population trans-ancestry PGS models to a three-population model, we found that the inclusion of Native Hawaiian GWAS continue to negatively impact the efficacy of the model (partial R^2^ = 0.034; Supplementary Figure S3).

**Figure 1.**
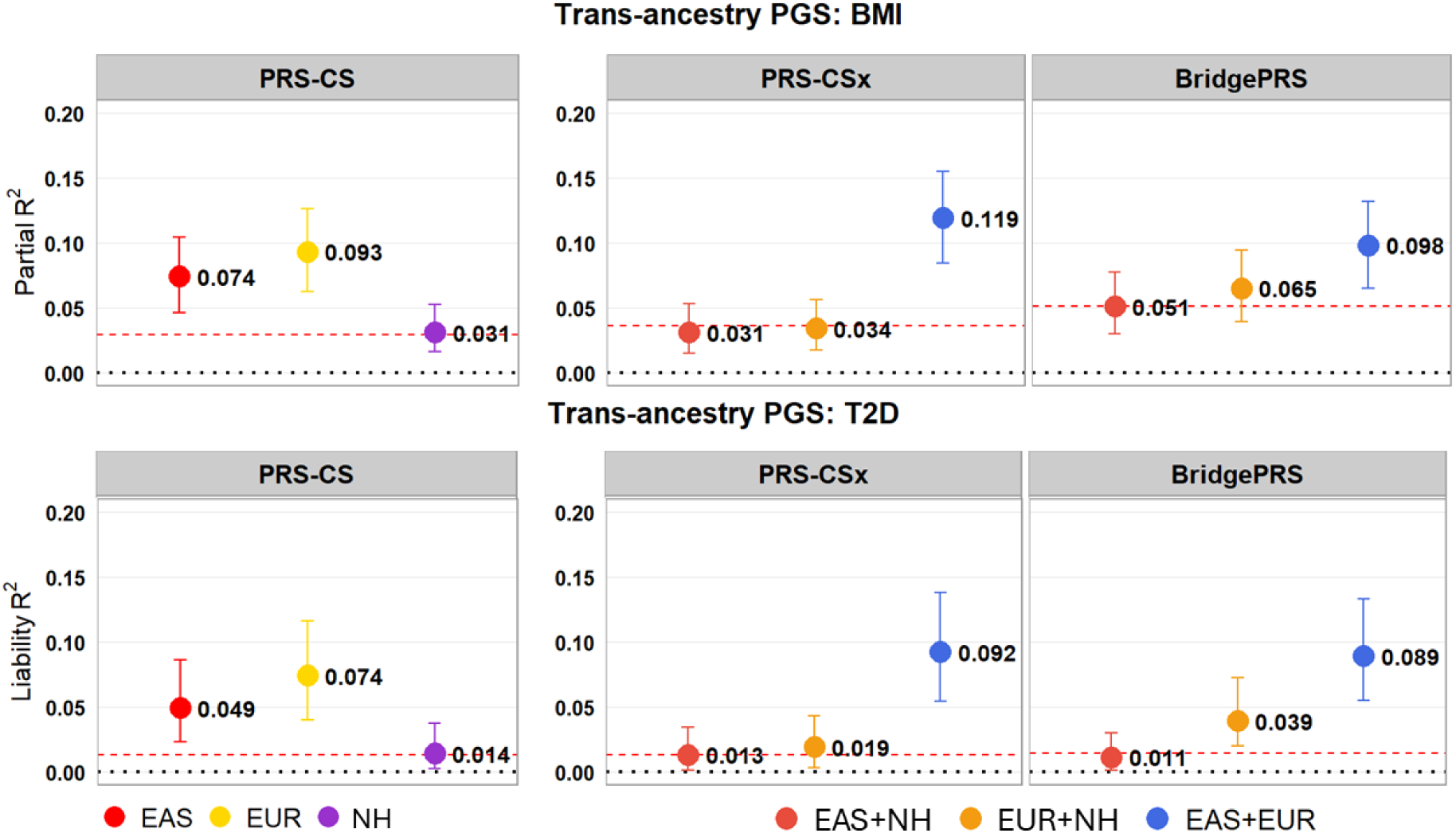
Trance-ethnic and single-population polygenic scores in Native Hawaiians. Multiple polygenic scores were estimated for each phenotype integrating information from single-population GWAS (European [EUR], East Asian [EAS], or Native Hawaiian [NH] alone) and combination of GWAS from two populations using either PRS-CSx or BridgePRS. (a) For BMI, the partial R^2^ represents the proportion of variance in BMI that is explained by PGS after adjusting for the first 10 PCs in validation dataset. (b) The liability R^2^ was used to evaluate the predictive ability of each PGS for T2D in Native Hawaiians after adjusting for age, sex, BMI, and first 10 PCs in validation data. Black dashed lines represent partial R^2^ and liability R^2^ of zero and red dotted lines crossing the lowest average variance explained among the three PGS models per each method.

Despite the improved performance of a trans-ancestry PGS combining EAS and EUR GWAS, because information from Native Hawaiian or Polynesian-ancestry cohorts could not be integrated effectively, we expect this model to continue to show the disparity in performance accuracy across population as previously observed for single-population PGS models (33). Indeed, we found the performance of these trans-ancestry PGS model to be the lowest among Native Hawaiians most enriched with Polynesian ancestry and among the Samoans (who are much less admixed) (**Figure 2**) (46).

**Figure 2.**
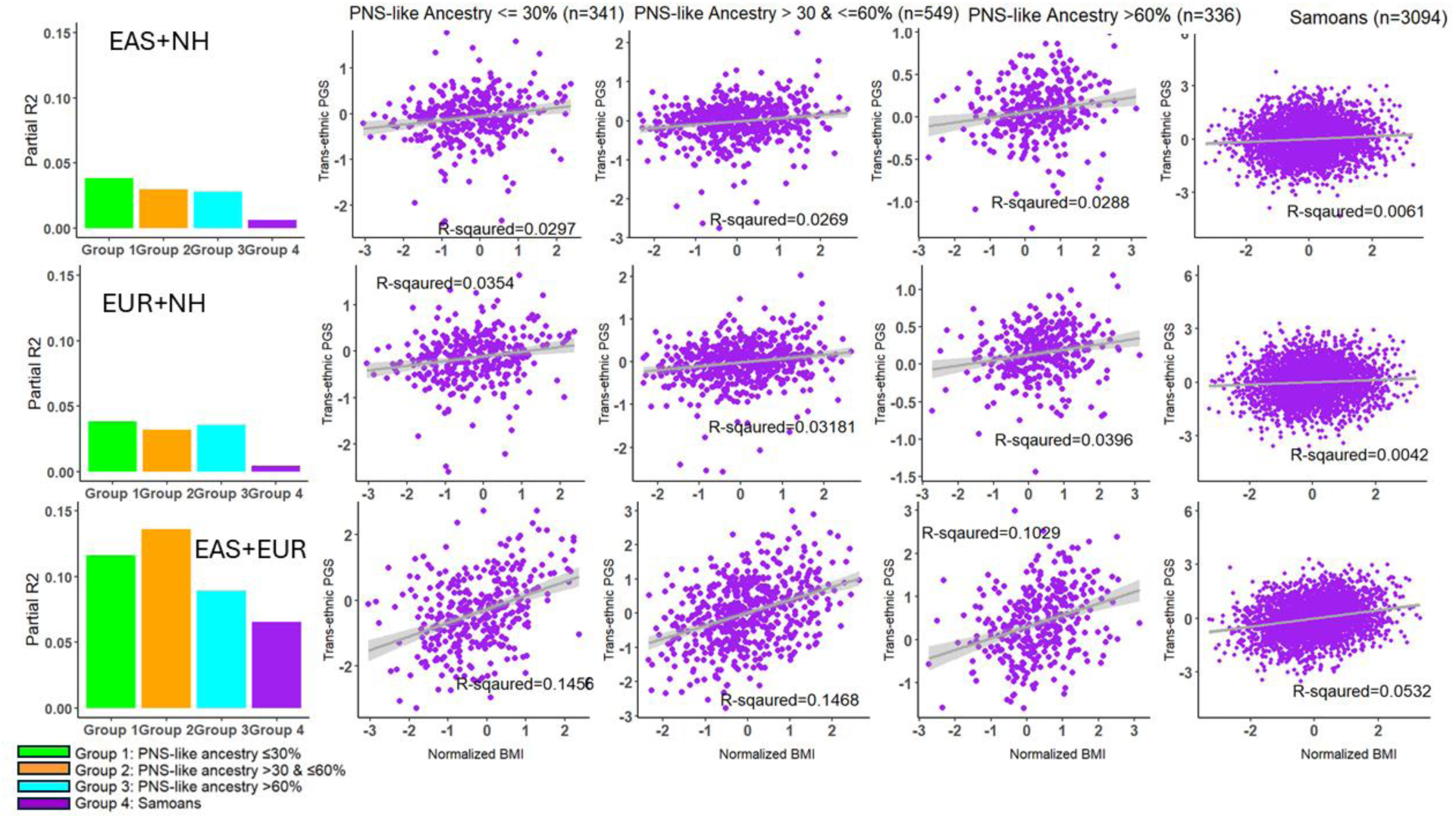
Performance of the trans-ethnic PGS models for BMI. Different approaches to combine GWAS summary statistics from multiple ethnic populations using PRS-CSx was evaluated and compared across strata by PNS-like ancestries. The PGS accuracy as measured by partial R^2^ was evaluated in different strata of Native Hawaiians based on estimated PNS-like ancestry proportions: < 30%, between 30% to 60%, and > 60%. Each PGS model was also validated in Samoans, who are taken as a largely-unadmixed representative of Western Polynesian ancestry.

For T2D, we observed much of the similar trend (**Figure 1**; Supplementary Table S2). A trans-ancestry PGS model combining information from EAS and EUR GWAS tend to perform better (liability R^2^ = 0.092) than single-population PGS (liability R^2^ = 0.014-0.074) model or trans-ancestry models including Native Hawaiians (liability R^2^ = 0.011-0.039), though in this case BridgePRS model slightly outperforms the PRS-CSx model (liability R^2^ = 0.090 vs. 0.092; **Figure 1**; Supplementary Table S2).

### Dietary characteristics across the Multiethnic Cohort

The correlation matrix (**Supplementary Figure S4, top**) displays relationships between the final list of 520 variables that were used to train our models. We provide the full list of these diet variables in supplement.

To visualize individual level variation in this high-dimensional space of dietary variables, we first applied principal components analysis (PCA) to the data after pruning the data for related variables (**Supplementary Figure S4, bottom**), and then projected the top 10 PCs down to two-dimensions with uniform manifold approximation and projection (UMAP). We performed this procedure across all five ethnic groups of MEC individuals (n = 191,183) to visualize and examine differential and shared dietary patterns between racial/ethnic groups. Nearly all Native Hawaiians in MEC were recruited from Hawaii and most of African-American and Latinos participants in MEC were recruited in Los Angeles. Japanese and Whites are more evenly distributed in terms of their recruitment locations (25). When we labeled individuals based on area of recruitment, it is clear that there may be shared food consumption behaviors among those recruited in Los Angeles (African Americans, Latinos, and some of the Whites), as well as those recruited in Hawai‘i (Native Hawaiians, Japanese, and some of the Whites), indicating a pattern perhaps driven by food type accessibility that differed by geographical regions (**Figure 3a**). A notable exception may be the Japanese in the MEC, who were more similar to each other regardless of the area they were recruited from, in contrast to, say, the Whites (**Figure 3a**), consistent with a stronger influence of consistent cultural practices in food choices.

**Figure 3.**
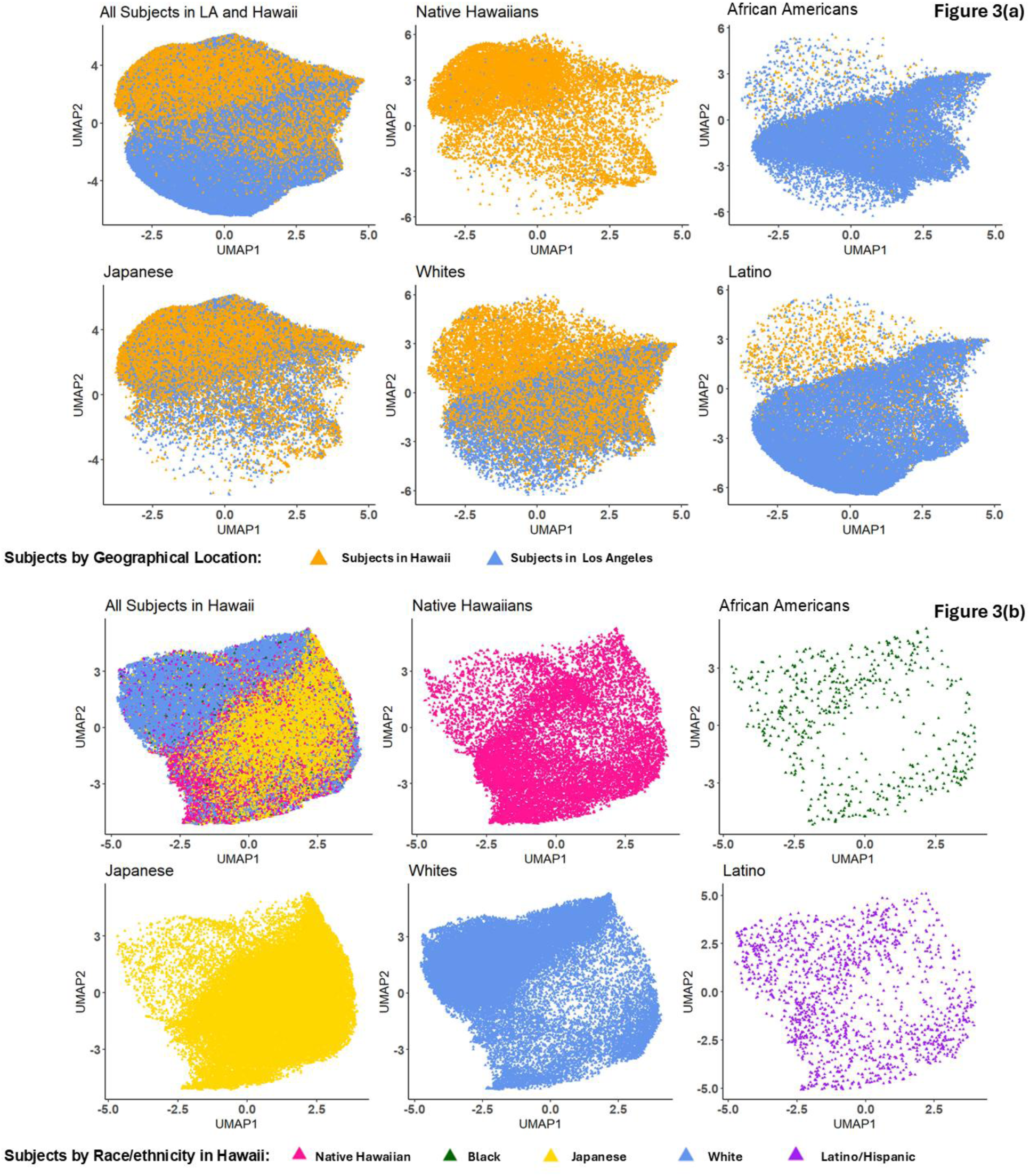
Dietary patterns/behaviors across MEC participants by geographical location and race/ethnicity. We applied uniform manifold approximation and projection (UMAP) to the top 10 principal components of the dietary data to visualize the diet/nutrition variability. UMAP is performed once across ethnic groups, but colored and visualized separately to aid visualization. UMAP was performed either (a) in all MEC participant, colored by recruitment area (Hawaii vs Los Angeles) or (b) in MEC participants recruited from Hawai‘I, colored by their self-identified race/ethnicity.

Restricting our analysis to only participants recruited from Hawai‘i so to lessen the influence of geographical constraints of food access, we repeated the UMAP analysis (**Figure 3b**). In this case, we observed that Japanese and Whites tend to differentiate on the 2-dimensional UMAP space, suggesting that ethnic- or culturally-driven influences on food choices. Furthermore, there may be shared food consumption behaviors among the Native Hawaiians with both the Japanese and Whites, as the pattern from Native Hawaiians span the UMAP space that overlapped with both the Japanese and Whites (**Figure 3b**). Indeed, by stratifying Native Hawaiians by estimated EAS-like or PNS-like ancestry, we found a closer affinity in UMAP space between Native Hawaiians with EAS-like ancestry and Japanese (Kullback-Leibler divergence, KLD_UMAP1_=0.08, KLD_UMAP2_=0.12) than the Native Hawaiians with PNS-like ancestry and Japanese (KLD_UMAP1_=0.23, KLD_UMAP2_=0.17; *P* < 0.0001 and *P* = 0.0004 by 10,000 rounds of bootstrapping, respectively) (**Supplementary Figure S5**). We observed an analogous pattern when stratified by EUR-like ancestry as well (*P* < 0.0001 for both UMAP1 and UMAP2 by bootstrapping; **Supplementary Figure S5**). Together, our findings suggest that there may be cultural effects on dietary choices, which is in part captured by genetic ancestry.

### Developing a risk stratification model based on dietary information for BMI and T2D

We used three different machine learning algorithms (**Methods**) to train and predict diet scores in Native Hawaiians (n=8,372 with survey data but no genetic data). Performance of these diet scores were evaluated for BMI and T2D in the held-out validation dataset (individuals with both survey and genetic data; n=5,223 for BMI, n=4,632 for T2D). Diet score models applying random forest algorithm outperformed score models trained by elastic net and neural network for both BMI and T2D (**Table 2**). For BMI, the model evaluation metrics suggest that a standard deviation (sd) increase in diet score was significantly associated with 0.38 sd (s.e of ±0.01) increase in BMI and explained approximately 12% of the variability after adjusting for daily energy intake (kcal/day) (**Table 2**). Moreover, a sd unit increase in T2D diet score was significantly associated with 21% increase in odds of having the disease after adjusting for age, sex, BMI, and daily energy intake (kcal/day). However, T2D diet scores estimated based on these three algorithms had relatively poor predictability (AUC ranged from 0.57 to 0.58) and explained relatively small variance of T2D, regardless of whether it is broadly defined incident T2D (Liability R^2^ ranged from 6.04×10^-3^ to 0.012), or incident T2D in 5 years from baseline (Liability R^2^ = 4.30×10^-3^; AUC = 0.54) (**Supplementary Table S3**).

**Table 2.**
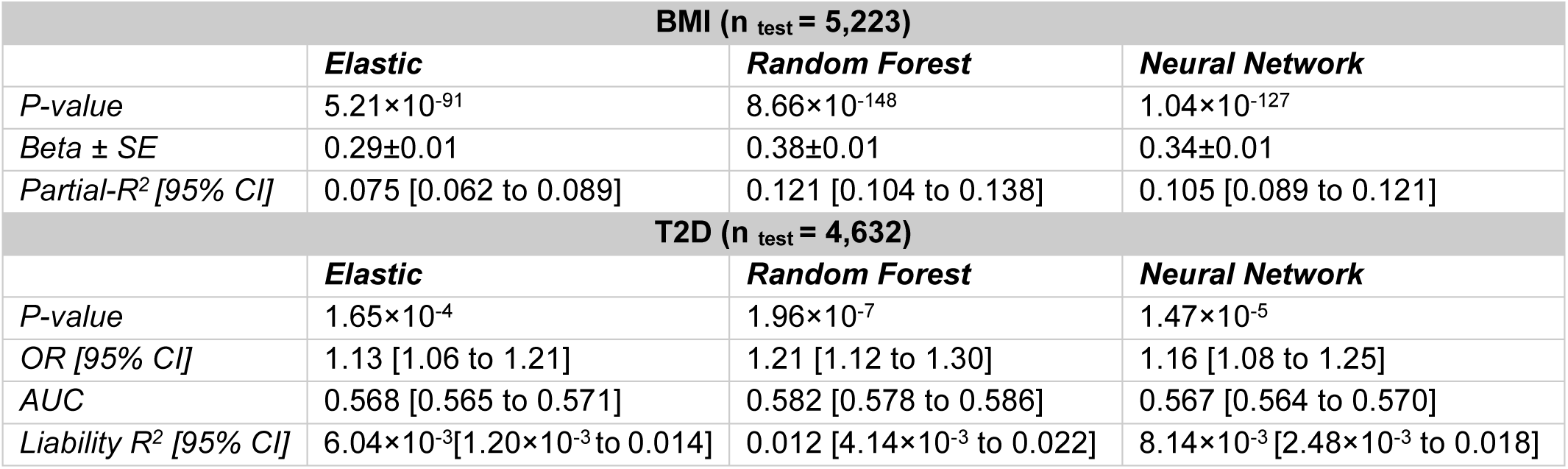
Diet scores trained and validated on BMI and T2D in Native Hawaiians. Performance of the predicted diet scores were evaluated and presented by phenotype and machine learning algorithm. Partial R^2^ for BMI was adjusted for daily energy intake (kcal/day). Odd ratio and liability R^2^ for T2D were adjusted for age, sex, BMI, and daily energy intake (kcal/day).

| **BMI (n<sub>test</sub> = 5,223)** |  |  |  |
| --- | --- | --- | --- |
|  | ***Elastic*** | ***Random Forest*** | ***Neural Network*** |
| *P-value* | $5.21 \times 10^{-91}$ | $8.66 \times 10^{-148}$ | $1.04 \times 10^{-127}$ |
| *Beta ± SE* | 0.29±0.01 | 0.38±0.01 | 0.34±0.01 |
| *Partial-R<sup>2</sup> [95% CI]* | 0.075 [0.062 to 0.089] | 0.121 [0.104 to 0.138] | 0.105 [0.089 to 0.121] |
| **T2D (n<sub>test</sub> = 4,632)** |  |  |  |
|  | ***Elastic*** | ***Random Forest*** | ***Neural Network*** |
| *P-value* | $1.65 \times 10^{-4}$ | $1.96 \times 10^{-7}$ | $1.47 \times 10^{-5}$ |
| *OR [95% CI]* | 1.13 [1.06 to 1.21] | 1.21 [1.12 to 1.30] | 1.16 [1.08 to 1.25] |
| *AUC* | 0.568 [0.565 to 0.571] | 0.582 [0.578 to 0.586] | 0.567 [0.564 to 0.570] |
| *Liability R<sup>2</sup> [95% CI]* | $6.04 \times 10^{-3}$ [$1.20 \times 10^{-3}$ to 0.014] | 0.012 [$4.14 \times 10^{-3}$ to 0.022] | $8.14 \times 10^{-3}$ [$2.48 \times 10^{-3}$ to 0.018] |

For the BMI, the diet score models developed here are much more strongly associate with BMI and explained much greater proportion of the variation in BMI than established dietary indices (**Supplementary Table S4**). This is perhaps not surprising as the diet score model was trained to specifically predict BMI. In the Native Hawaiian cohort, we also find that the diet scores are significantly associated with future BMI, taken approximately 10 years after baseline (n=3,492; partial R^2^ [95%CI] = 0.12 [0.10 to 0.14]), though we note that adult BMI tend to be stable over time. While T2D diet scores generally explained little of the phenotypic variation on the liability scale, it still appeared to be more strongly associated with the outcome than other dietary indices, including outperforming empirical lifestyle or dietary indices for insulin resistance (**Supplementary Table S5**).

Finally, we examined how well the diet score models trained and validated in Native Hawaiians would perform in other MEC populations, focusing on the BMI diet score model as it appeared to be most efficacious. As is the case with PGS models, we also found that diet score models generally performed less well in explaining variation in BMI in other ethnic groups, particularly for Latinos (partial R^2^ = 0.027 – 0.056 across diet scores; **Supplementary Figure S6**). Our results thus suggest that diet score models currently are not well-transferred across populations, even if the same level of dietary information is available, likely because our diet scores were trained on Native Hawaiian diet data while the dietary patterns differ across ethnic groups (**Figure 3**).

### Combining PGS and diet score in risk stratification model

As we expect the genetic and non-genetic predictors would capture complementary information in the variation of the phenotype of interest, we fitted regression models with the best-performing trans-ancestry PGS and diet score as the main predictors on each of BMI and T2D to assess their individual and joint effects adjusting for potential confounding variables (**Table 3 and 4**). For BMI, the multivariate model containing both PGS and diet score (along with other covariates) is a significantly better predictor than models with either of the scores (adjusted R^2^ in Model 3 = 0.29 vs. adjusted R^2^ in Model 1 or 2 = 0.21; *P* = 4.95×10^-117^ and *P* = 4.89×10^-120^, respectively by likelihood ratio test; **Table 3.a and 3.b**), suggesting that adding diet score to PGS model significantly improved the model fit. Both the PGS and diet scores capture complementary and nearly additive information, as the partial R^2^ barely diminished in the joint model (0.12 or 0.11 in Models 1 or 2, respectively, to 0.10 in Model 3). We did not observe a significant interaction between PGS and diet score, whether treating the scores as continuous variables (Model 4) or as categorical variables (Model 5 and 6), though the effect coefficients of diet score in Model 5 and 6 may hint at a non-linear effect with upper quartiles than the first quartile of the diet score (**Table 3.a**).

**Table 3.** Regression models incorporating both polygenic score and diet score in Native Hawaiians for BMI. a). Relationship and predictive power of both PGS and diet score were assessed by estimating their individual effects on BMI as the main predictor or jointly in the same model while controlling for covariates and potential confounding (smoking status, physical activity, education level, daily energy intake (kcal/day) and first 10 PCs). Diet variable was treated as both continuous and categorical variables. Potential interaction effects of PGS and diet score on BMI was also evaluated by adding a multiplicative interaction term in regression model; b). Model comparisons were performed using the log-likelihood ratio test. *ΔR²* indicates the changes in adjusted R2 between the more complex model to the simpler model.

| **(a). BMI** |  |  |  |  |  |
| --- | --- | --- | --- | --- | --- |
| **n=5,225** | **Variables** | **Beta** | **P-value** | **Partial R<sup>2</sup>** | **Adjusted R<sup>2</sup>** |
| Model 1 | PGS | 0.39 | 6.80 x10<sup>-134</sup> | 0.12 | 0.21 |
| Model 2 | Diet Score | 0.36 | 1.36 x10<sup>-131</sup> | 0.11 | 0.21 |
| Model 3 | PGS | 0.35 | 3.98 x10<sup>-119</sup> | 0.10 | 0.29 |
|  | Diet Score | 0.32 | 5.12 x10<sup>-116</sup> | 0.10 |  |
| Model 4 | PGS | 0.35 | 4.20 x10<sup>-119</sup> | 0.10 | 0.29 |
|  | Diet Score | 0.32 | 5.46 x10<sup>-116</sup> | 0.10 |  |
|  | PGS :Diet Score | -6.25 | 0.96 | 5.53 x10<sup>-7</sup> |  |
| Model 5 | PGS | 0.36 | 2.41 x10<sup>-120</sup> | 0.10 | 0.28 |
|  | Diet Score Q2 | 0.27 | 1.89 x10<sup>-14</sup> | 1.18 x10<sup>-2</sup> |  |
|  | Diet Score Q3 | 0.51 | 5.35 x10<sup>-47</sup> | 4.12 x10<sup>-2</sup> |  |
|  | Diet Score Q4 | 0.77 | 3.69 x10<sup>-88</sup> | 7.73 x10<sup>-2</sup> |  |
| Model 6 | PGS | 0.36 | 3.88 x10<sup>-42</sup> | 0.04 | 0.28 |
|  | Diet Score Q2 | 0.27 | 3.94 x10<sup>-14</sup> | 0.01 |  |
|  | Diet Score Q3 | 0.51 | 2.59 x10<sup>-46</sup> | 0.04 |  |
|  | Diet Score Q4 | 0.77 | 1.50 x10<sup>-86</sup> | 0.08 |  |
|  | PGS : Diet Score Q2 | 1.66x10<sup>-3</sup> | 0.96 | 4.76 x10<sup>-7</sup> |  |
|  | PGS : Diet Score Q3 | -1.22x10<sup>-2</sup> | 0.73 | 2.49 x10<sup>-5</sup> |  |
|  | PGS : Diet Score Q4 | -3.12x10<sup>-3</sup> | 0.93 | 1.64 x10<sup>-6</sup> |  |

**Table 3.** Regression models incorporating both polygenic score and diet score in Native Hawaiians for BMI.
| **(b). BMI** |  |  |  |  |  |
| --- | --- | --- | --- | --- | --- |
| **Comparison** | **Models Compared** | **df** | **χ<sup>2</sup>** | **p-value** | **Δ R<sup>2</sup>** |
| Model 1 vs Model3 | PGS vs PGS + diet | 1 | 528.88 | 4.95x10<sup>-117</sup> | 0.0795 |
| Model 2 vs Model3 | Diet vs PGS + diet | 1 | 542.69 | 4.89x10<sup>-120</sup> | 0.0817 |
| Model 1 vs Model4 | PGS vs PGS + diet + PGS×diet | 2 | 528.89 | 1.42x10<sup>-115</sup> | 0.0795 |
| Model 2 vs Model4 | Diet vs PGS + diet + PGS×diet | 2 | 542.70 | 1.42x10<sup>-118</sup> | 0.0817 |
| Model 3 vs Model4 | Additive vs Interaction | 1 | 0.0085 | 0.93 | 1.21x10<sup>-6</sup> |
| Model 1 vs Model5 | PGS vs PGS + diet_Qs | 3 | 438.77 | 8.83x10<sup>-95</sup> | 0.0665 |
| Model 2 vs Model5 | Diet vs PGS + diet_Qs | 3 | 452.59 | 8.97x10<sup>-98</sup> | 0.0688 |
| Model 1 vs Model6 | PGS vs PGS + diet_Qs + PGS×diet_Qs | 6 | 438.98 | 1.16x10<sup>-91</sup> | 0.0666 |
| Model 2 vs Model6 | Diet vs PGS + diet_Qs + PGS×diet_Qs | 6 | 452.79 | 1.23x10<sup>-94</sup> | 0.0688 |
| Model 4 vs Model6 | Continuous vs Categorical | 4 | 89.91 | 1.37x10<sup>-18</sup> | -0.0129 |
| Model 5 vs Model6 | Additive vs Interaction | 3 | 0.2045 | 0.98 | 2.97x10<sup>-5</sup> |

For T2D, the PGS appeared to have a stronger association and better predictability than diet score based on OR, liability R^2^ or AUC as an independent predictor and when included in the same model with diet score (**Table 4.a**). The log-likelihood ratio tests suggest that the joint effects of PGS and diet score in Model 3 statistically improved the model fit compared to single predictor models, though the improvement was marginal but significant (*ΔR^2^ **=***0.0028 for Model 1 vs 3 and *ΔR^2^ **=***0.032 for Model 2 vs 3 shown in **Table 4.b**). Similar trends were observed when diet score was treated as a categorical variable. The odds of having T2D increased by 16%, 23%, and 52% for the 2^nd^, 3^rd^, and 4^th^ quartiles, respectively, when compared to the 1^ST^ quartile (Model 5 in **Table 4.a**). Interactions between PGS and diet score (whether modeled as continuous and categorical) were observed; the interaction between two appears to decrease the odds of having T2D.

**Table 4.** Logistic regression models for polygenic risk score and diet score in Native Hawaiians on T2D. a). The relationship and predictive power of both PGS and diet score were assessed by estimating their individual effects on T2D as the main predictor or independent variable and as a covariate to each other controlling for other covariates and potential confounding. Diet variable was treated as both continuous and categorical variables. The combined effect of PGS and diet score on T2D was evaluated by adding a multiplicative interaction term in logistic regression model; b). Each logistic regression model was compared to each other applying the log-likelihood test. All models for T2D were adjusted for age, sex, BMI, smoking status, physical activity, education level, daily energy intake (kcal/day), and first 10 PCs.

| **(a). T2D** |  |  |  |  |  |
| --- | --- | --- | --- | --- | --- |
| **n=4,383** | **Variables** | **OR [95% CI]** | **P-value** | **Liability R<sup>2</sup> [95% CI]** | **AUC** |
| Model 1 | PGS | 1.71 [1.59 to 1.86] | 4.29 x10<sup>-41</sup> | 0.0830 [0.061 to 0.108] | 0.7295 |
| Model 2 | Diet Score | 1.17 [1.09 to 1.26] | 3.19 x10<sup>-5</sup> | 0.008 [0.002 to 0.017] | 0.7042 |
| Model 3 | PGS | 1.72 [1.59 to 1.86] | 5.19 x10<sup>-41</sup> | 0.0825 [0.060 to 0.107] | 0.7312 |
|  | Diet Score | 1.03 [1.02 to 1.03] | 3.81 x10<sup>-5</sup> |  |  |
| Model 4 | PGS | 1.72 [1.59 to 1.87] | 3.02 x10<sup>-41</sup> | 0.083 [0.060 to 0.107] | 0.7322 |
|  | Diet Score | 1.18 [1.09 to 1.27] | 3.73 x10<sup>-5</sup> |  |  |
|  | PGS :Diet Score | 0.92 [0.85 to 0.98] | 1.75 x10<sup>-2</sup> |  |  |
| Model 5 | PGS | 1.72 [1.59 to 1.86] | 2.64 x10<sup>-41</sup> | 0.083 [0.061 to 0.108] | 0.7312 |
|  | Diet Score Q2 | 1.16 [0.96 to 1.40] | 0.12 |  |  |
|  | Diet Score Q3 | 1.23 [1.02 to 1.49] | 0.03 |  |  |
|  | Diet Score Q4 | 1.52 [1.23 to 1.88] | 8.26 x10<sup>-5</sup> |  |  |
| Model 6 | PGS | 2.04 [1.79 to 2.38] | 3.17 x10<sup>-19</sup> | 0.038 [0.026 to 0.054] | 0.7320 |
|  | Diet Score Q2 | 1.15 [0.95 to 1.38] | 0.14 |  |  |
|  | Diet Score Q3 | 1.22 [1.01 to 1.48] | 0.04 |  |  |
|  | Diet Score Q4 | 1.55 [1.27 to 1.91] | 1.00 x10<sup>-4</sup> |  |  |
|  | PGS : Diet Score Q2 | 0.83 [0.68 to 1.02] | 0.06 |  |  |
|  | PGS : Diet Score Q3 | 0.79 [0.65 to 0.97] | 0.02 |  |  |
|  | PGS : Diet Score Q4 | 0.84 [0.68 to 1.03] | 0.04 |  |  |

**Table 4.** Logistic regression models for polygenic risk score and diet score in Native Hawaiians on T2D.
| **(b). T2D** |  |  |  |  |  |
| --- | --- | --- | --- | --- | --- |
| **Comparison** | **Models Compared** | **df** | **$\chi^2$** | **p-value** | **$\Delta R^2$** |
| Model 1 vs Model3 | PGS vs PGS + diet | 1 | 16.89 | 3.97x10<sup>-5</sup> | 0.0028 |
| Model 2 vs Model3 | Diet vs PGS + diet | 1 | 192.79 | 7.83x10<sup>-44</sup> | 0.0324 |
| Model 1 vs Model4 | PGS vs PGS + diet + PGS×diet | 2 | 22.71 | 1.17x10<sup>-5</sup> | 0.0038 |
| Model 2 vs Model4 | Diet vs PGS + diet + PGS×diet | 2 | 198.61 | 7.44x10<sup>-44</sup> | 0.0334 |
| Model 3 vs Model4 | Additive vs Interaction | 1 | 5.83 | 0.0158 | 0.001 |
| Model 1 vs Model5 | PGS vs PGS + diet_Qs | 3 | 15.92 | 0.0012 | 0.0027 |
| Model 2 vs Model5 | Diet vs PGS + diet_Qs | 3 | 191.82 | 2.47x10<sup>-41</sup> | 0.0323 |
| Model 1 vs Model6 | PGS vs PGS + diet_Qs + PGS×diet_Qs | 6 | 23.09 | 7.68x10<sup>-4</sup> | 0.0039 |
| Model 2 vs Model6 | Diet vs PGS + diet_Qs + PGS×diet_Qs | 6 | 198.99 | 3.11x10<sup>-40</sup> | 0.0335 |
| Model 4 vs Model6 | Continuous vs Categorical | 4 | 0.38 | 0.9844 | 1.00 x10<sup>-4</sup> |
| Model 5 vs Model6 | Additive vs Interaction | 3 | 7.17 | 0.0667 | 0.0012 |

### Prediction of chronic diseases using BMI diet score (mediation analysis)

Because BMI is a risk factor for a number of chronic diseases, we tested the association of the BMI diet score on the incidence of multiple chronic diseases and conditions in MEC participants. We found that the BMI diet score was associated with 21% increase in odds of developing T2D, 20% increase in odds of having stroke and chronic kidney disease, 17% higher odds of having ischemic heart disease, and 12% increase in having anemia (**Table 5, Supplementary Figure S7**). These associations were found even after adjusting BMI directly in the model, suggesting that the effect due to the diet might be mediated through other mechanisms in addition to BMI. We conducted mediation analysis for each of these five diseases with four different biomarkers that are available to us in the cohort, including CRP (as an inflammatory marker), HDL, LDL, and Triglyceride (reflecting different types of lipids) (**Supplementary Table S6**). While after correcting for a total of 20 tests we do not generally have significant findings (Bonferroni *p-*value threshold = 0.0025), we did observe several suggestive associations for mediation effects. For example, the average causal mediation effect (ACME) of CRP on T2D (ACME =0.0022, p-value=0.06) suggests that the BMI-related diet could lead to incident T2D through the inflammatory pathway. We also found that BMI-related diet may affect the levels of healthy lipids (HDL) in the system that could play a role in developing chronic kidney disease (ACME=0.0028, p-value=0.02) or stroke (ACME=0.0020, p-value=0.06). For T2D, approximately 13% of the overall relationship between BMI-associated diet and phenotype was attributed by triglycerides (p-value=0.04); there were 23% increased odds of T2D for each unit increase in triglycerides (p-value=4.35×10^-10^).

**Table 5.** The association of BMI diet score with other disease endpoints. Only diseases with significant association after correcting for multiple testing burden (0.05 / 24 traits = 2.1x10^-3^) were shown. Anemia was included as its p-value was very close to the threshold. For the complete list of results please refer to **Supplementary Figure S7**.

| Phenotype | P-value | N<sub>case</sub> | N<sub>control</sub> | OR [95% CI] |
| --- | --- | --- | --- | --- |
| **Anemia** | $3.76 \times 10^{-3}$ | 1760 | 1800 | 1.12 [1.03 to 1.21] |
| **Chronic Kidney Disease** | $3.01 \times 10^{-6}$ | 1556 | 1989 | 1.20 [1.12 to 1.31] |
| **Ischemic Heart Disease** | $1.33 \times 10^{-4}$ | 1627 | 1944 | 1.17 [1.08 to 1.27] |
| **Stroke** | $7.14 \times 10^{-4}$ | 561 | 2921 | 1.20 [1.08 to 1.34] |
| **T2D** | $1.17 \times 10^{-7}$ | 1898 | 2734 | 1.21 [1.13 to 1.30] |

## DISCUSSION

In the present study we developed comprehensive risk stratification models integrating genetic and dietary predictors for BMI and type-2 diabetes in Native Hawaiians. Our trans-ancestry polygenic score approach showed improved prediction accuracy over previous single-ancestry models for Native Hawaiians (15), demonstrating that leveraging large-scale cross-ancestry GWAS summary statistics can improve prediction accuracy in populations with limited genetic studies. Beyond genetic factors, dietary intake emerged as a powerful complementary predictor, with our hypothesis-free diet-based models explaining substantial variance in BMI. The combination of polygenic scores and dietary predictors achieved markedly improved prediction performance compared to either approach alone. Furthermore, our best-performing BMI diet score showed associations with multiple chronic diseases, even when BMI was included as a covariate, suggesting that BMI-associated dietary patterns may influence broader health outcomes through inflammatory and lipid-mediated pathways.

### Comparative predictive power of diet versus genetics

Building on these predictive capabilities, our analysis revealed that dietary intake explains phenotypic variation beyond genetic factors. Our best-performing diet score accounted for approximately 12% of adult BMI variance, demonstrating substantially higher explanatory power than Mediterranean diet scores (∼0.6-3%), Healthy Eating Index (∼3.9%), and other established dietary pattern indices (1-3%) (**Supplementary Table S4**) (47–50). In contrast, the best-performing diet model trained on T2D incidence, though significantly associated with the outcome in held-out validation cohort, explained only ∼1% of phenotypic variation on the liability scale. This limited predictive ability for T2D likely reflects several factors: dietary patterns vary considerably over time, making past diet a poor predictor of future disease onset; and individuals with pre-diabetic conditions may have already modified their diets toward healthier patterns, confounding the diet-disease relationship.

Interpreting the major contributing factors for a machine learning-based prediction model is a general challenge (51–52). In our best performing approach (Random Forest), we can obtain features of importance, but it lacks directionality. ElasticNet model, while did not attain as high of an accuracy, it does provide a list of variables and direction of effects in association with T2D incidence or BMI. We found that the effect sizes of elastic net-selected dietary variables were generally relatively small for T2D incidence compared to BMI (**Supplement Data**). As expected, there were shared dietary patterns between BMI and T2D given that BMI is an established T2D risk factor. For example, we observed the effect of established risk factors including higher red meat consumption and diet soda intake were positively associated with both BMI and T2D (53–55). Interestingly, we also observed that more non-Western dietary patterns (ramen/saimin, refried beans, enchiladas) were associated with reduced T2D, which also had suggestive association previously in literature (56). Taken together, population-specific dietary-metabolic relationships warrant further investigation.

### Mechanisms linking diet to chronic disease risk

Despite the challenges to establish causality with diet scores, we found BMI diet score to be strongly associated with incidence of some chronic disease status in Native Hawaiians, such as T2D, chronic kidney disease, ischemic heart disease and stroke (Table 5). These associations were observed even after adjusting for BMI, suggesting that dietary effects involve mechanisms independent of body weight. To begin probing the mechanism through which BMI-related diet is associated with incident chronic diseases, we turned to mediation analysis. Our analysis revealed that the biomarkers (CRP, HDL, LDL, Triglycerides) we tested explain less than 13% of the association between the dietary scores and chronic disease risk, indicating that the majority of diet’s health effects operate through other molecular markers in the inflammatory or lipid metabolism pathways, or through other unmeasured biological pathways. However, we also note that our mediation analysis is preliminary and limited given the relatively small number of individuals with available molecular biomarkers.

Future studies could consider emerging pathways such as gut microbiome interactions, metabolomic alterations, epigenetic mechanisms, and direct cellular mechanisms (including mitochondrial and endothelial function) to capture the full impact of diet on chronic disease risk (57–61). Population-specific mechanisms should also be investigated, such as genetic variants affecting metabolic enzyme function, population-specific genetic adaptations in Native Hawaiians, and traditional versus Western diet interactions (62–65). Nevertheless, due to this apparent ’pleiotropic’ effect of dietary patterns associated with BMI, these BMI-prone diet scores could be incorporated into clinical risk assessment tools to improve chronic disease risk prediction.

### Gene-diet interactions and the CREBRF paradox

In our combined risk stratification models, we found that both genetics and dietary patterns significantly and independently influence metabolic health, together significantly improving regression model fits for both BMI and T2D. Interestingly, for T2D, we observed a counterintuitive negative gene-diet interaction that contributed to reduced disease odds. This unexpected finding may reflect a critical limitation of trans-ancestry approaches: while our trans-ancestry PGS models based on European and East Asian GWAS outperformed other models overall, they systematically missed the single most important population-specific variant with strong metabolic effects in Native Hawaiians—the CREBRF variant (64).

CREBRF was strongly associated with both BMI and T2D in Samoans and Native Hawaiians (66–67). When we added the CREBRF variant to our combined risk stratification model (containing both trans-ancestry PGS model and diet score model), including this single variant significantly improved model fit and explained approximately 0.7% and 0.2% more variation in BMI and T2D, respectively (**Supplementary Table S7).** In contrast, incorporating Native Hawaiian GWAS data as additional ancestry when training the trans-ancestry PGS models resulted in poor performance and appeared to add noise due to small sample size (**Figure 1, Supplementary Table S2**). The observed T2D gene-diet interaction could represents a ’missing heritability’ problem and highlights the limitations of PGS portability across populations. Indeed, given the small cohort sizes that are likely insurmountable in genetic epidemiology studies of diverse or indigenous populations, further improvement for polygenic risk predictions may require leveraging both trans-ancestry PGS models from other continental ancestries as well as population-enriched variants identified in the population of interests.

Moreover, CREBRF is uniquely paradoxical: it increases BMI while simultaneously protecting against T2D. This challenges the conventional causal pathway where high BMI or obesity as a major risk factor for T2D development and suggests different endophenotypes within both BMI and diabetes, indicating heterogeneity in BMI-diabetes relationships (68–69). Additionally, recent admixture in Native Hawaiians may interact differently with this variant under different environmental conditions, adding further complexity (63). This has important clinical implications: some Native Hawaiians carrying this variant may be obese or overweight but metabolically healthy and may not face elevated T2D risk. Further investigations on the CREBRF paradox are required to better understand the underlying mechanisms that could help reduce the high prevalence of obesity and T2D in Native Hawaiians.

### Limitations and future directions

One current limitation of our diet score models is that it is dependent on variable availability, which differs across cohorts. Unlike genetic data which is more standardized or can be imputed to the same set of genetic markers for calculating the polygenic scores, our diet score models currently require having the same set of diet information available to calculate the score. Indeed, we have examined the possibility of harmonizing the available diet phenotype in the MEC with that from UK Biobank through manual curation but found that variables can only match ∼30-57% of the times depending on the strictness of interpretation. Therefore, to utilize diet scores in risk stratification in different cohorts will require extensive and systematic variable harmonization of dietary surveys or assessment tools.

Another limitation is that while our composite scores effectively predict metabolic phenotypes, establishing a causal role of diet remains challenging. Unlike polygenic scores, which are determined at conception and remain constant throughout life, dietary information in the MEC was collected in adulthood, when participants may already have been in the preliminary stages of disease but not yet diagnosed. As a step toward establishing causality, we tested whether our best-performing BMI diet score could predict BMI measured approximately 10 years after baseline, finding significant predictive ability (n=3492; partial R^2^ [95%CI] = 0.12 [0.10 to 0.14]), although we should note that adult BMI is generally stable and our two measurements of BMI, 10 years apart, are highly correlated (R^2^ = 0.736). Future longitudinal study designs that collect detailed dietary data, particularly in adolescent or young adults, could greatly benefit efforts to establish causal relationships between diet and metabolic conditions (70–73).

### Implications for genetic studies and clinical practice

Irrespective of the clinical utilities of the diet score models, at least for BMI and to a lesser extent for T2D, it is clear that diet information explains part of the phenotypic variation that are not captured by the best available polygenic predictors. This has important implications for genetic research: when diet information is available for a cohort in a genetic study, computing and adjusting for diet scores will help remove a part of the phenotypic variation explained by the non-genetic factors, thereby improving the statistical power of the genetic mapping study. Such an increase in power will be particularly valuable when mapping population-specific associations in a relatively small GWAS cohort, as is often the case for underrepresented populations like Native Hawaiians.

## CONCLUSION

For populations with limited genetic data, integrating large-scale GWAS data from other ancestries substantially improved polygenic score performance compared to single-ancestry approaches in Native Hawaiians. Similarly, data-driven diet score models outperformed established dietary indices in explaining BMI variance (12% vs. 0.6-3.9%). Importantly, diet scores trained on shared risk factors such as BMI can be applied across different chronic diseases, though effectiveness may vary by the disease, the population and dietary patterns. Population-enriched genetic mechanisms such as the CREBRF paradox we observed, underscore the critical need for conducting genetic and dietary research within diverse populations rather than relying solely on findings from populations of European ancestry. Our findings demonstrate that personalized metabolic disease risk assessment should integrate both genetic and dietary information, accounting for population-specific genetic variants and dietary patterns to develop effective prevention and management strategies for diverse populations.

## Data Availability

All data produced in the present study are available upon reasonable request to the authors

## ACKNOWLEDGEMENT

We would also like to thank all of the research participants in the Multiethnic Cohort that are involved in this study. The Multiethnic Cohort was funded through grants from the National Cancer Institute (U01CA164973, P01CA168530) and the National Human Genome Research Institute (U01HG007397). This study is supported by grants from the National Human Genome Research Institute (R01HG011646 to C.W.K.C.). We would also like to acknowledge the involvement of the Samoan Obesity, Lifestyle, and Genetic Adaptations (OLaGA) Study Group (some of whom are listed co-authors):

*Ranjan Deka*, Dept of Environmental Health, University of Cincinnati, Cincinnati, OH, USA *Nicola L. Hawley*, Dept of Chronic Disease Epidemiology, Yale University, New Haven, CT, USA *Stephen T. McGarvey*, Dept of Epidemiology and International Health Institute, School of Public Health, and Dept of Anthropology, Brown University, Providence, RI, USA *Ryan L. Minster*, Dept of Human Genetics, University of Pittsburgh, Pittsburgh, PA, USA *Take Naseri*, International Health Institute, School of Public Health, Brown University, Providence, RI, USA & Naseri Associates Public Health Consultancy Firm and Family Health Clinic, Apia, Samoa *Satupa‘itea Viali*, Faculty of Medicine, National University of Samoa, Apia, Samoa, and Dept of Chronic Disease Epidemiology, Yale University, New Haven, CT, USA *Muagututi‘a Sefuiva Reupena*, Lutia i Puava ‘ae Mapu I Fagalele, Apia, Samoa *Daniel E. Weeks*, Dept of Human Genetics and Biostatistics & Health Data Science, University of Pittsburgh, Pittsburgh, PA, USA Finally, computation for this work was supported by the University of Southern California’s Center for Advanced Research Computing (https://www.carc.usc.edu/).

---

**Figure S1.**
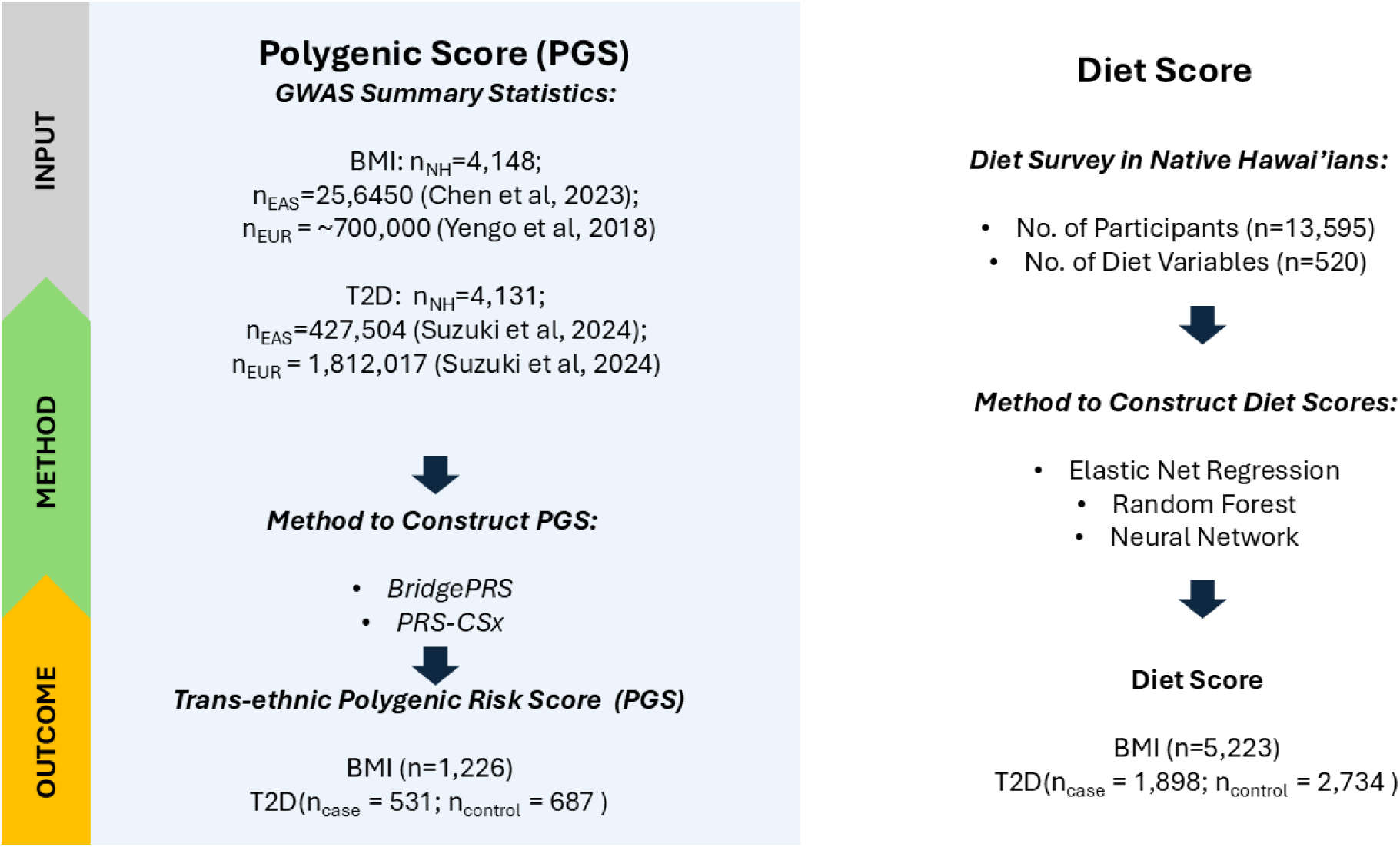
Overview for constructing and evaluating trans-ethnic polygenic score (PGS) and diet score prediction models. The estimations of trans-ethnic PGS for BMI and T2D were based on the GWAS from three ethnic groups including East Asian (EAS, representing largely EAS-like ancestry), European (EUR, representing largely EUR-like ancestry), and Native Hawaiians (NH, representing largely Polynesian, or PNS-like, ancestry) using Bayesian approaches (BridgePRS and PRS-CSx). In this case, MEC-NH individuals genotyped on MEGA array (N = 4,131-4,148) were used to train the PGS, while individuals genotyped on the GDA array (N = 1,218 – 1,226) were used as validation dataset. Diet score was trained and validated for each phenotype using three different machine learning-based algorithms, with all individuals with survey data only (N = 7026_T2D_; 8372_BMI_) used as training dataset, and individuals with survey and genetic data (N = 4632 _T2D_ ; 5,223 _BMI_) were used as validation dataset.

**Figure S2.**
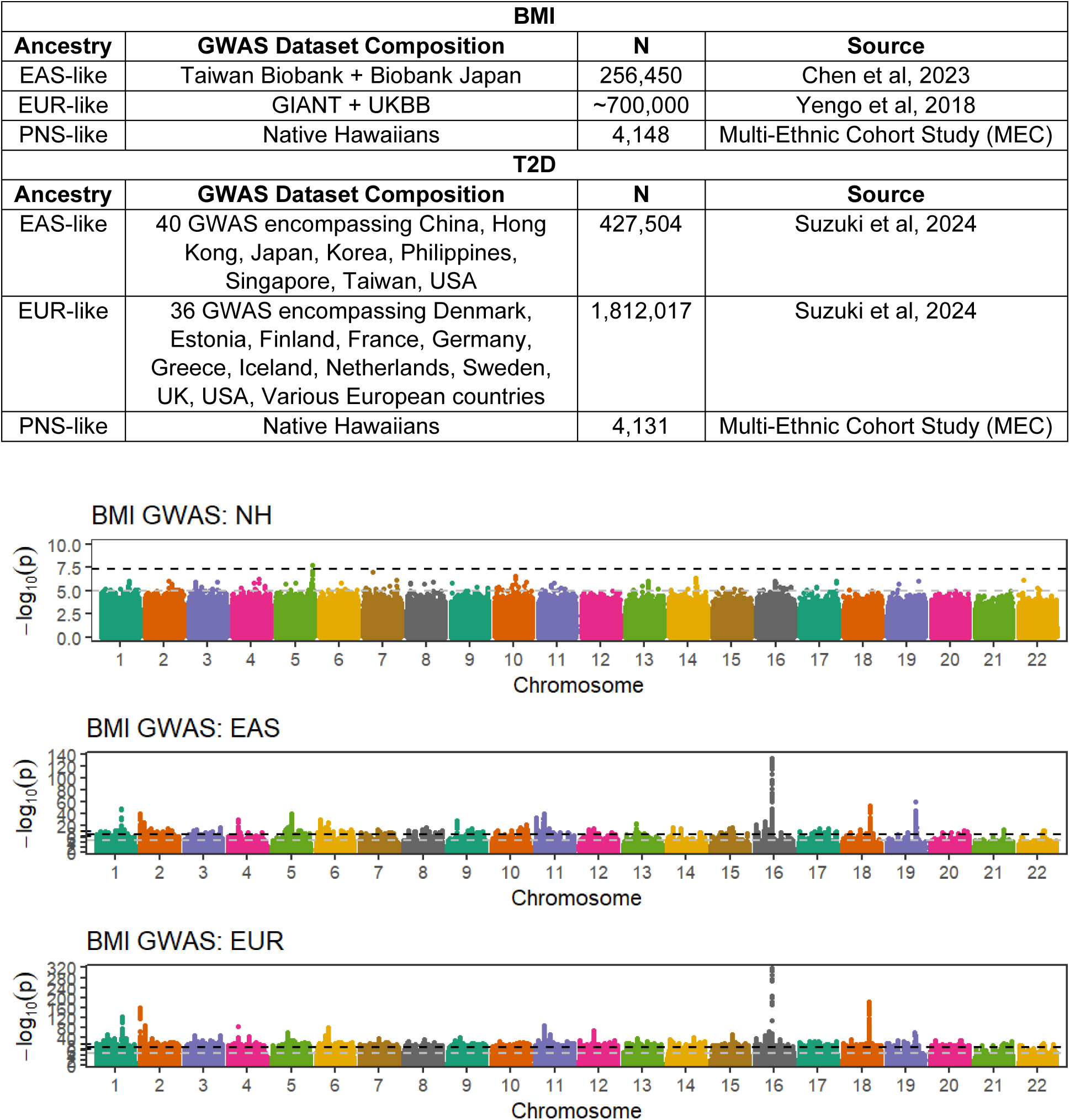
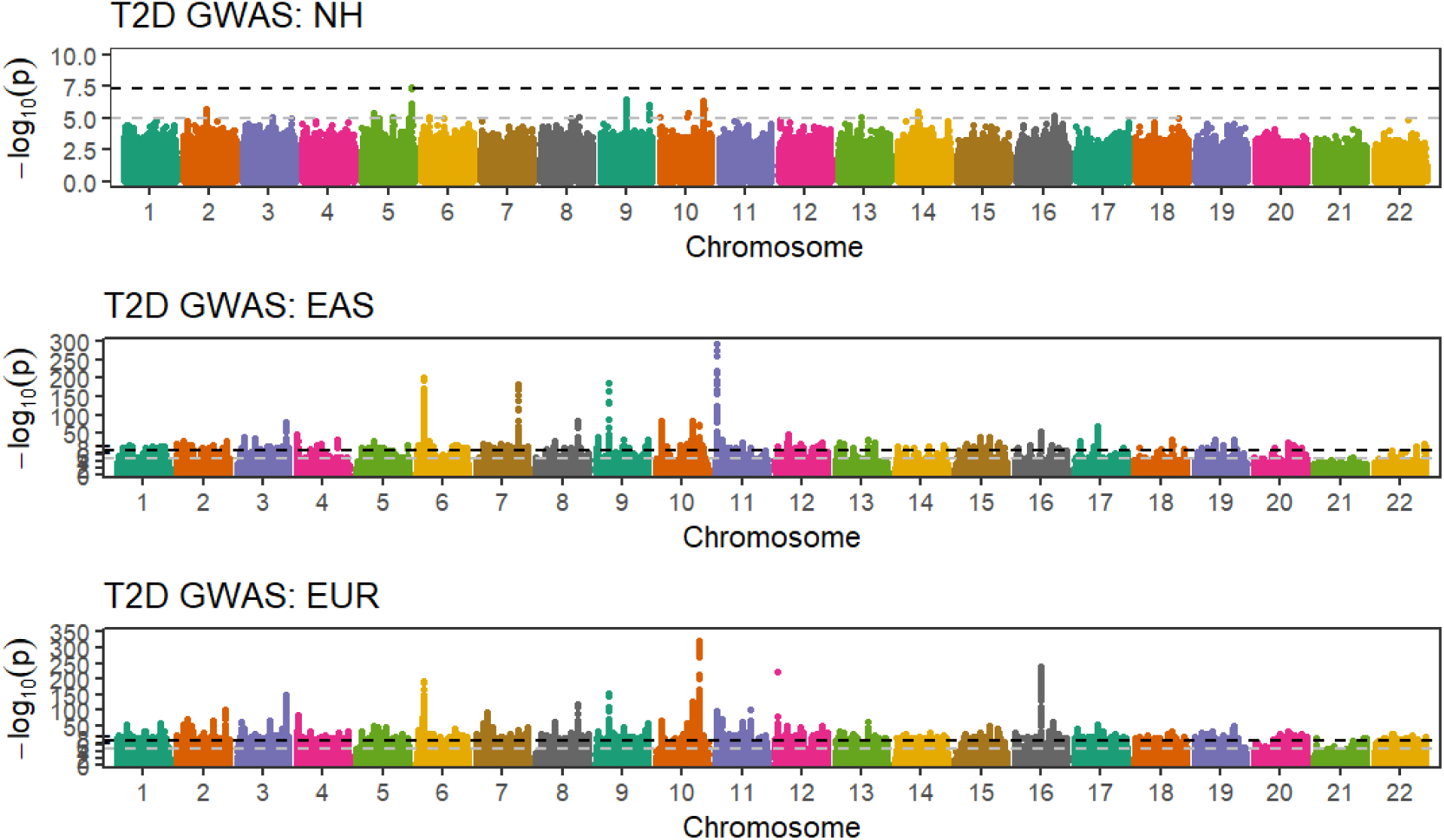
Summary of GWAS datasets that were used to train PGS models for BMI (top) and T2D (bottom) for Native Hawaiians.

**Figure S3.**
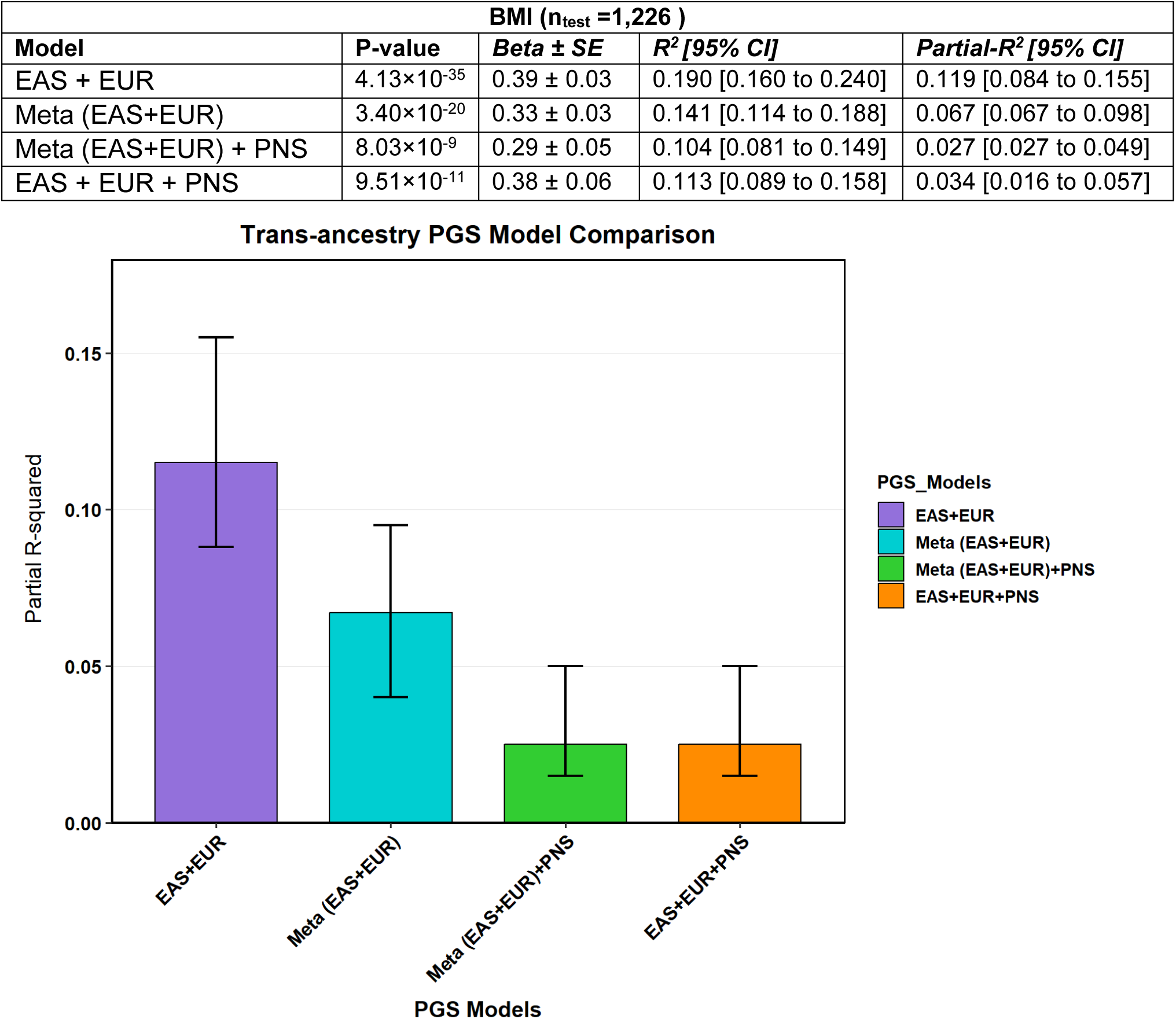
Comparison of trans-ethnic PGS models integrating GWAS information from two and three ancestral groups using PRS-CSx. We compared two additional approaches for constructing PGS prediction models for BMI: (1) Meta (EAS+EUR), which performs GWAS meta-analysis and then apply PRS-CS on the single GWAS summary statistics, and (2) Meta (EAS+EUR) + PNS, which we combined the meta-analyzed summary statistics from (1) with that from PNS-only for a trans-ethnic PGS model using PRS-CSx. This approach could in theory incorporate information from three separate populations.

**Figure S4.**
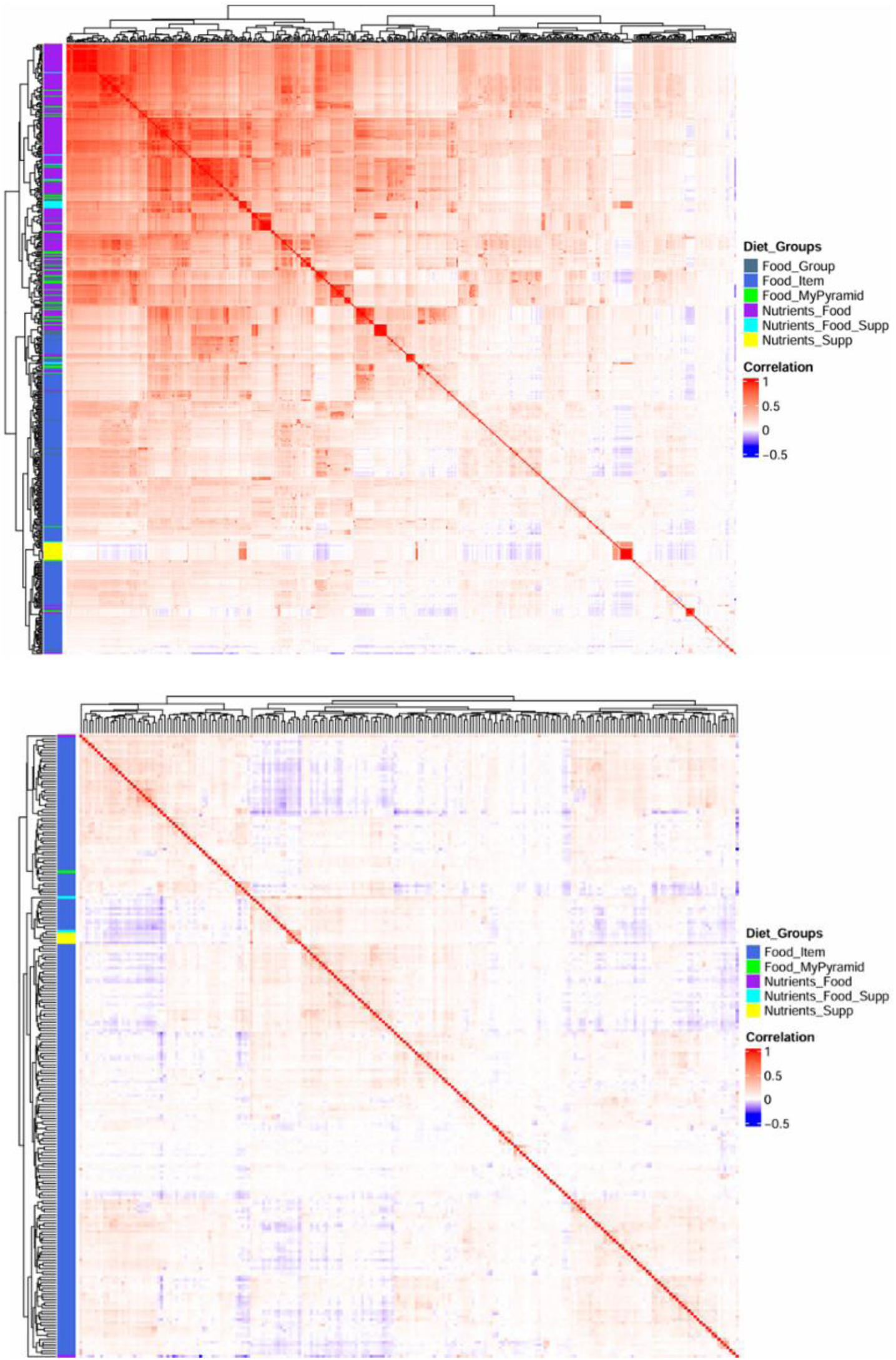
Correlation among dietary variables available in the Multi-Ethnic Cohort. The correlations among (top) all 520 diet variables and (bottom) pruned (n=220) diet variables in Native Hawaiians (N = 13,595). The pruned dataset removes randomly one variable among all pairs of variables correlated with R^2^ > 0.5.

**Figure S5.**
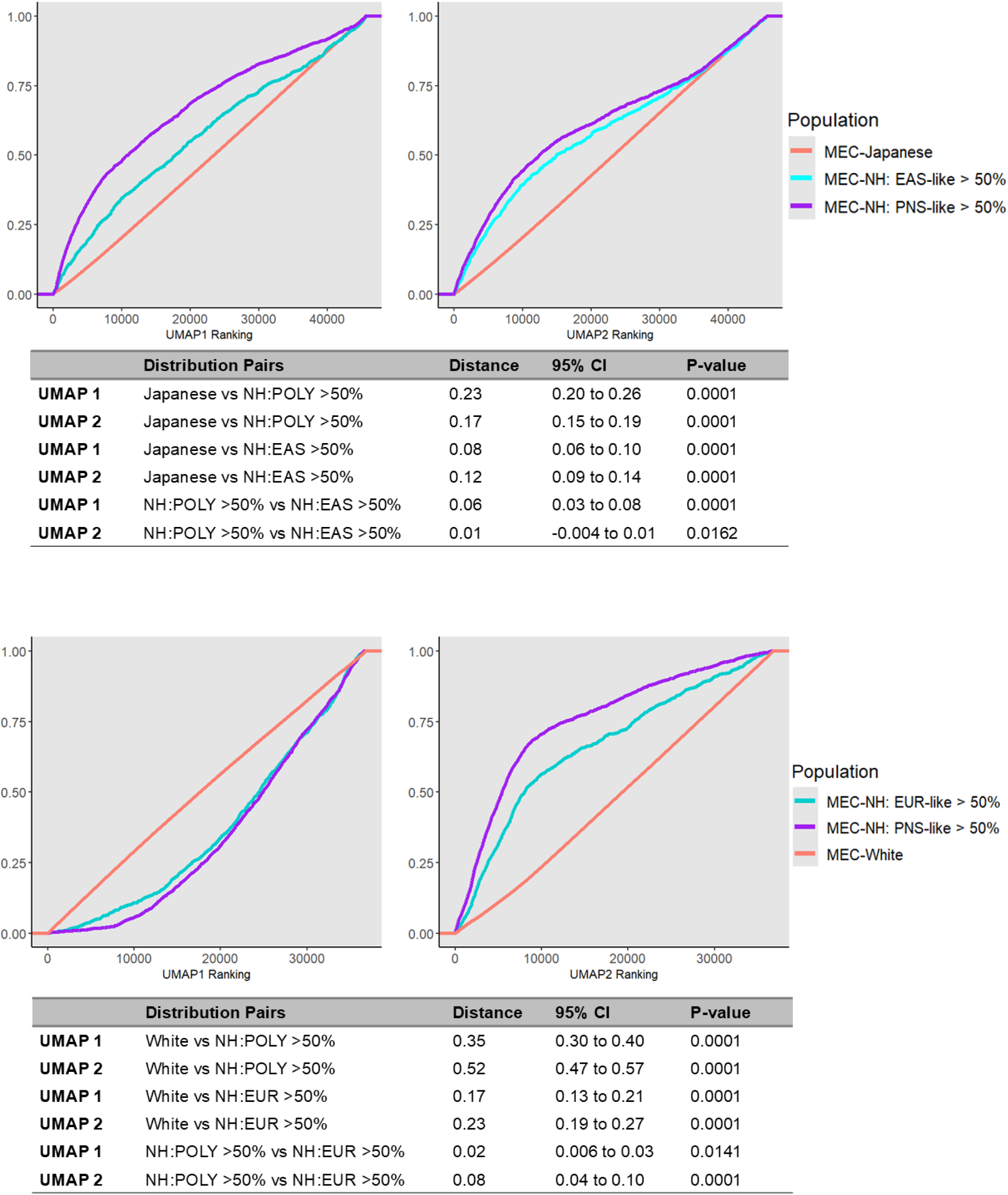
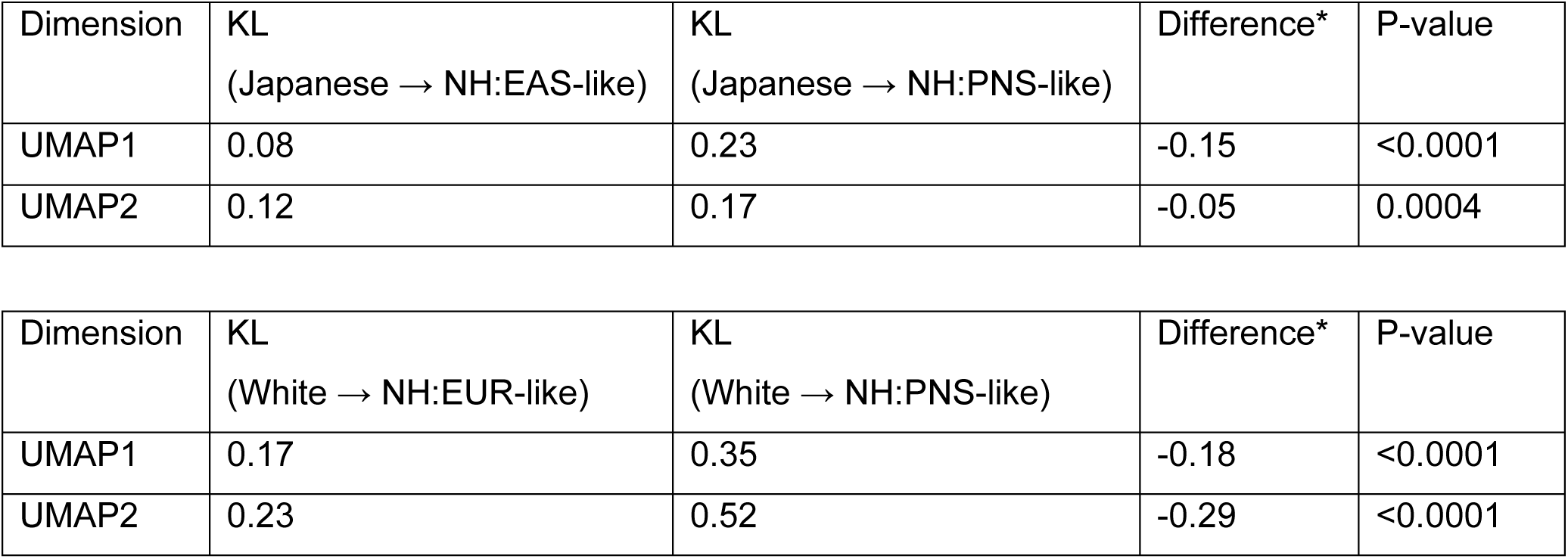
Similarity in dietary patterns by genetic ancestries in Native Hawaiians. Stratifying Native Hawaiians by their estimated majority genetic ancestry, we computed the Kullback-Leibler (KL) divergence as a distance metric, using either the MEC-Japanese or MEC-White as the reference. For each rank ordered UMAP dimension, we first calculated the observed KL divergences from the Japanese reference population to NH:EAS-like (Native Hawaiians with EAS-like ancestry >50%) and to NH:PNS-like (Native Hawaiians with PNS-like ancestry > 50%) separately using bootstrap estimation with 100 iterations (kld_ci_bootstrap function from the kldest R package). The observed difference was then computed as KL(Japanese || NH:EAS) - KL(Japanese || NH:POLY). To determine whether this observed difference could arise by chance, we performed 10,000 rounds of permutation of the majority ancestry label in Native Hawaiians. We recalculated the differences in KL divergences for each of the 10,000 rounds of permutation to obtain the empirical p-value. This procedure was repeated for UMAP2 dimension and for comparisons to MEC-Whites as reference population.

**Figure S6.**
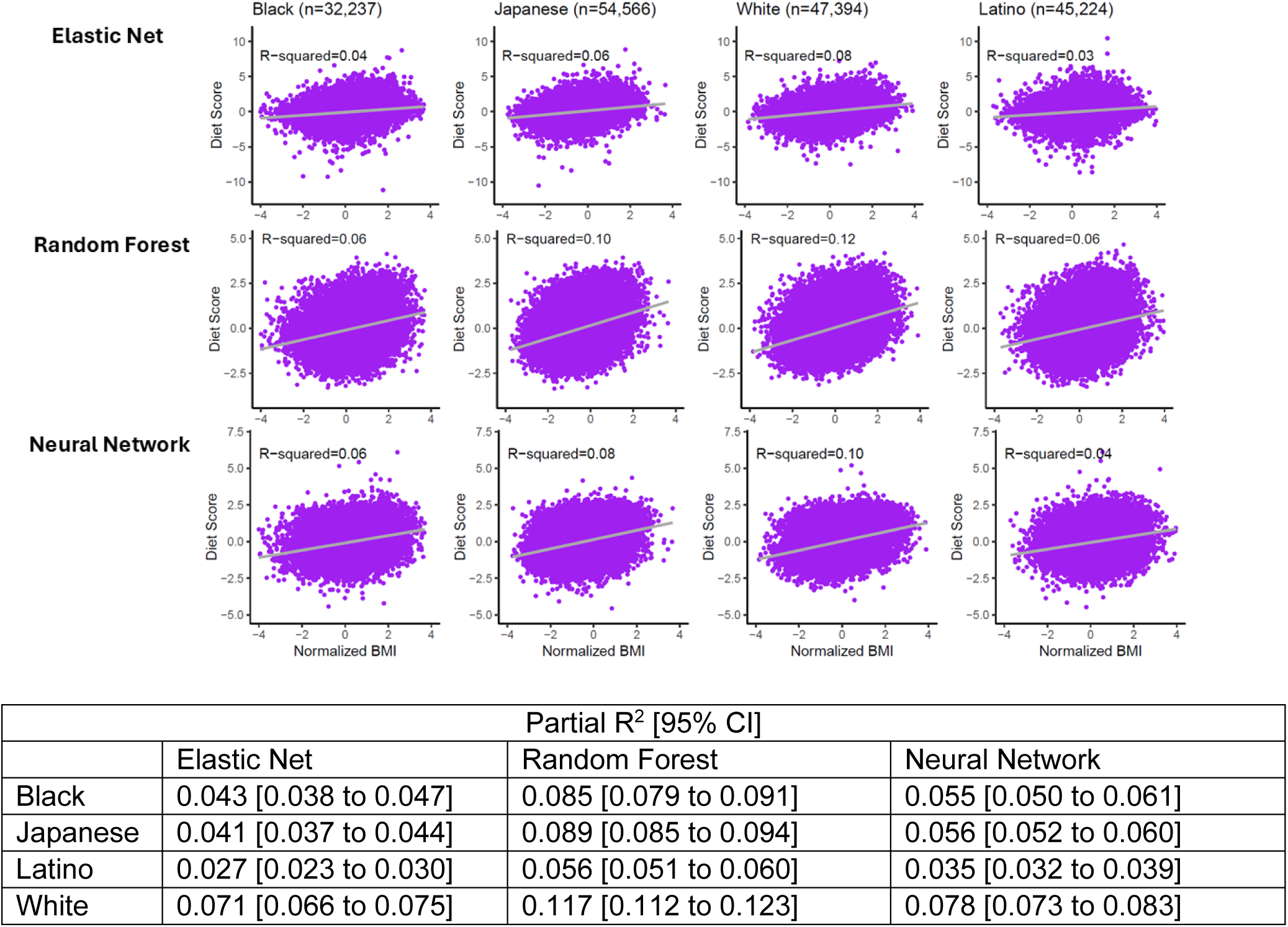
Transferability of diet scores for BMI across other racial/ethnic groups in MEC participants. The partial R^2^ for each diet score, based on a model adjusted for daily calorie intake is also shown in the table.

**Figure S7.**
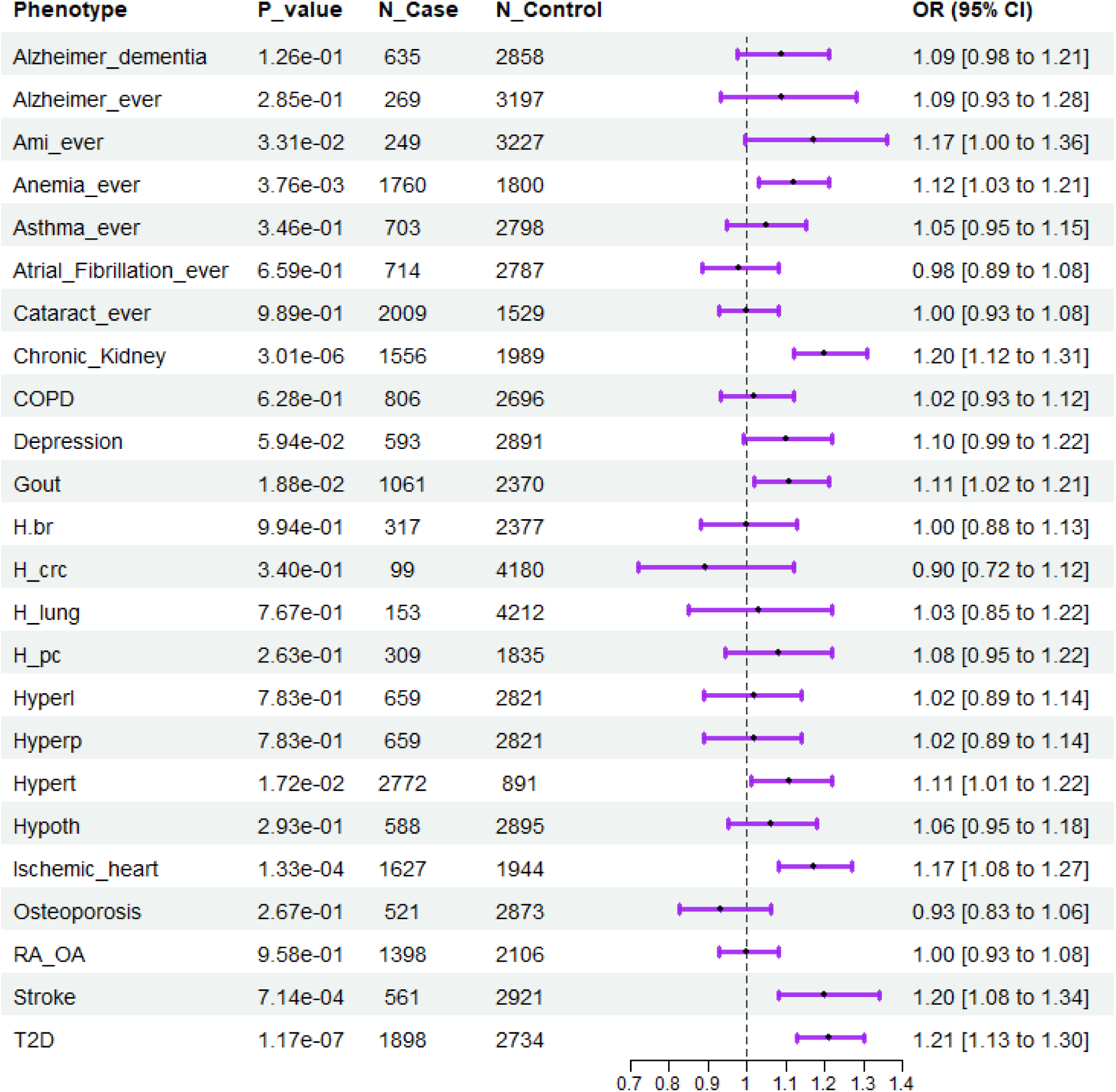
Association of BMI diet score with other binary disease outcome in Native Hawaiians. For each disease, cases were defined based on ICD-9 and ICD-10 codes in the Medicare claim data, following the Chronic Condition Warehouse database (**Methods**). The remaining individuals are assumed to be controls. The only exception is T2D, which was defined separately for this study (**Methods**). The associations were assessed after adjusting for age, sex, BMI, and first 10 PCs.

**Table S1.**
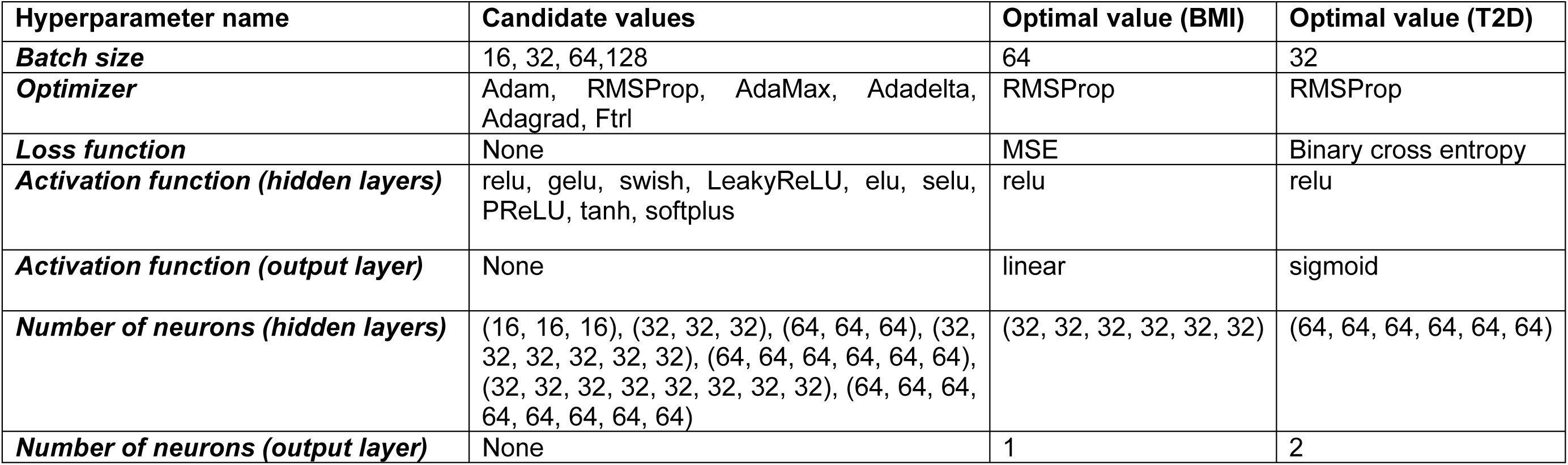
The candidate and optimal values of the hyperparameters of the neural network models for predicting BMI and T2D. Column Optimal value (BMI) and Optimal value (T2D) show the values of the hyperparameters in the models with the lowest validation loss for predicting BMI and T2D, respectively. None in Column Candidate values indicates that the corresponding optimal hyperparameters were determined by following common practices. For example, for predicting binary-valued T2D, the activation function and number of neurons at the output layer are commonly set to Binary cross-entropy and 2, respectively.

**Table S2.** PGS accuracy on BMI and T2D across different PGS models. All BMI PGS were adjusted for the first 10 PCs. T2D PGS were adjusted for the age, sex, BMI, and first 10 PCs. PGS models were validated in held-out sample of Native Hawaiians that were not used in generating the GWAS dataset.

| BMI (n<sub>test</sub> =1226 ) |  |  |  |  | T2D (n<sub>test</sub> =1218 ) |  |  |  |
| --- | --- | --- | --- | --- | --- | --- | --- | --- |
| *Model* | *GWAS* | *P-value* | *Beta ± SE* | *Partial-R<sup>2</sup> [95% CI]* | *OR* | *P-value* | *AUC* | *Liability R<sup>2</sup> [95% CI]* |
| *PRS-CS* | *EAS* | 6.00×10<sup>-22</sup> | 0.26 ± 0.03 | 0.074 [0.046 to 0.104] | 2.15 [1.70 to 2.89] | 2.54×10<sup>-8</sup> | 0.52 | 0.049 [0.023 to 0.086] |
|  | *EUR* | 1.26×10<sup>-27</sup> | 0.40 ± 0.04 | 0.093 [0.062 to 0.126] | 2.59 [2.02 to 3.53] | 1.16×10<sup>-11</sup> | 0.63 | 0.074 [0.040 to 0.116] |
|  | *PNS* | 2.68×10<sup>-9</sup> | 0.33 ± 0.05 | 0.029 [0.014 to 0.051] | 1.56 [1.17 to 2.09] | 2.55×10<sup>-3</sup> | 0.56 | 0.014 [0.002 to 0.037] |
| *PRS-CSx* | *EAS+PNS* | 1.61×10<sup>-13</sup> | 0.39 ± 0.05 | 0.044 [0.024 to 0.069] | 1.43 [1.12 to 1.85] | 7.69×10<sup>-3</sup> | 0.53 | 0.011 [0.001 to 0.030] |
|  | *EUR+PNS* | 2.12×10<sup>-11</sup> | 0.33 ± 0.05 | 0.036 [0.019 to 0.061] | 1.71 [1.41 to 2.13] | 6.31×10<sup>-7</sup> | 0.62 | 0.039 [0.015 to 0.072] |
|  | *EAS+EUR* | 4.13×10<sup>-35</sup> | 0.39 ± 0.03 | 0.119 [0.084 to 0.155] | 2.29 [1.90 to 2.86] | 7.80×10<sup>-14</sup> | 0.64 | 0.090 [0.055 to 0.133] |
| *BridgePRS* | *EAS+PNS* | 1.69×10<sup>-15</sup> | 0.22 ± 0.03 | 0.051 [0.030 to 0.077] | 1.20 [1.06 to 1.35] | 3.87×10<sup>-3</sup> | 0.56 | 0.013 [0.001 to 0.034] |
|  | *EUR+PNS* | 2.31×10<sup>-19</sup> | 0.25 ± 0.03 | 0.065 [0.039 to 0.094] | 1.26 [1.10 to 1.43] | 5.41×10<sup>-4</sup> | 0.58 | 0.019 [0.003 to 0.043] |
|  | *EAS+EUR* | 7.16×10<sup>-29</sup> | 0.35 ± 0.03 | 0.098 [0.065 to 0.132] | 1.78 [1.55 to 2.11] | 4.68×10<sup>-14</sup> | 0.65 | 0.092 [0.054 to 0.138] |

**Table S3.** Association between diet score and incident T2D. Diet score associations with incident type 2 diabetes within 5 years from the baseline in Native Hawaiians (n=535). The diet score was trained using random forest. Odd ratios (OR) and liability R^2^ for T2D were adjusted for age, sex, BMI, and daily energy intake (kcal/day). AUC= area under the receiver operating characteristic curve.

| **Model** | **OR (95%CI)** | **P-value** | **AUC** | **Liability R<sup>2</sup> (95%CI)** |
| --- | --- | --- | --- | --- |
| T2D incidence in ≤5 yrs (n=535) | 0.94 (0.82 to 1.03) | 0.28 | 0.54 | 0.0043 (2.71×10<sup>-5</sup> to 0.03) |

**Table S4.** Association of dietary scores and indices to BMI in Native Hawaiians. Evaluations were conducted only in held-out Native Hawaiians that were not used to develop the diet score in this study. The associations between dietary scores/indices and BMI were adjusted for calorie intake. We did not test the empirical lifestyle index for hyperinsulinemia and empirical lifestyle index for insulin resistance as these two indices used directly BMI as part of their model construction.

| BMI |  |  |  |  |
| --- | --- | --- | --- | --- |
|  | N | P-value | Beta ± SE | Partial-R<sup>2</sup> [95% CI] |
| *Elastic Net* | 5222 | 3.62×10<sup>-81</sup> | 0.27 ± 0.01 | 0.068 [0.054 to 0.081] |
| *Random Forest* | 5222 | 1.36×10<sup>-142</sup> | 0.37 ± 0.01 | 0.117 [0.100 to 0.134] |
| *Neural Network* | 5222 | 2.17×10<sup>-115</sup> | 0.32 ± 0.01 | 0.0952 [0.0800 to 0.1110] |
| *HEI total score* | 5224 | 4.70×10<sup>-5</sup> | -0.05 ± 0.01 | 0.0032 [0.0009 to 0.0071] |
| *AHEI total score* | 5224 | 0.0020 | -0.04 ± 0.01 | 0.0018 [0.0002 to 0.0049] |
| *DASH total score* | 5224 | 1.90×10<sup>-7</sup> | -0.07 ± 0.01 | 0.0052 [0.0020 to 0.0101] |
| *AMDS E total score* | 5224 | 1.44×10<sup>-7</sup> | -0.07 ± 0.01 | 0.0053 [0.0022 to 0.0101] |
| *AMDS total score* | 5224 | 3.87×10<sup>-6</sup> | -0.07 ± 0.02 | 0.0041 [0.0015 to 0.0080] |
| Overall plant-based diet index | 5224 | 7.50×10<sup>-12</sup> | -0.09 ± 0.01 | 0.0090 [0.0044 to 0.0148] |
| Healthful plant-based diet index | 5224 | 5.37×10<sup>-12</sup> | -0.09 ± 0.01 | 0.0091 [0.0048 to 0.0148] |
| Unhealthful plant-based diet index | 5224 | 0.5000 | 0.01 ± 0.01 | 8.74×10<sup>-5</sup> [2.52×10<sup>-7</sup> to 0.0015] |
| Empirical dietary index for hyperinsulinemia | 5069 | 1.12×10<sup>-28</sup> | 0.16 ± 0.01 | 0.0234 [0.0151 to 0.0331] |
| Empirical dietary inflammatory pattern | 5069 | 5.67×10<sup>-37</sup> | 0.17 ± 0.01 | 0.0305 [0.0210 to 0.0421] |
| Empirical dietary index for insulin resistance | 5224 | 4.26×10<sup>-43</sup> | 0.20 ± 0.01 | 0.0357 [0.0264 to 0.0471] |
| HEI-2015: Total vegetables | 5224 | 0.7140 | -0.0049 ± 0.01 | 2.58×10<sup>-5</sup> [1.12×10<sup>-7</sup> to 0.0012] |
| HEI-2015: Greens & beans | 5224 | 0.0013 | -0.04 ± 0.01 | 0.0020 [0.0004 to 0.0050] |
| HEI-2015: Total fruits | 5224 | 0.0003 | -0.05 ± 0.01 | 0.0025 [0.0005 to 0.0063] |
| HEI-2015: Whole fruits | 5224 | 0.1760 | -0.02 ± 0.01 | 0.0004 [1.42×10<sup>-6</sup> to 0.0023] |
| HEI-2015: Whole grains | 5224 | 0.0021 | -0.04 ± 0.01 | 0.0018 [0.0002 to 0.0048] |
| HEI-2015: Dairy | 5224 | 0.4410 | -0.01 ± 0.02 | 0.0001 [4.79×10<sup>-7</sup> to 0.0015] |
| HEI-2015: Total protein foods | 5224 | $2.82 \times 10^{-8}$ | $0.08 \pm 0.01$ | 0.0059 [0.0024 to 0.0103] |
| HEI-2015: Seafood & plant proteins | 5224 | 0.8580 | $0.0026 \pm 0.01$ | $6.13 \times 10^{-6}$ [$1.13 \times 10^{-7}$ to 0.0010] |
| HEI-2015: Fatty acids | 5224 | 0.1730 | $0.02 \pm 0.02$ | 0.0004 [$1.96 \times 10^{-6}$ to 0.0019] |
| HEI-2015: Sodium | 5224 | $4.75 \times 10^{-12}$ | $-0.09 \pm 0.01$ | 0.0091 [0.0043 to 0.0145] |
| HEI-2015: Refined grains | 5224 | $8.77 \times 10^{-14}$ | $-0.10 \pm 0.01$ | 0.0106 [0.0056 to 0.0168] |
| HEI-2015: Saturated fats | 5224 | 0.0103 | $-0.04 \pm 0.02$ | 0.0013 [$7.69 \times 10^{-5}$ to 0.0040] |
| HEI-2015: Added sugars | 5224 | 0.0052 | $0.04 \pm 0.01$ | 0.0015 [$9.94 \times 10^{-5}$ to 0.0042] |
| Healthy Eating Index-2015: Total score | 5224 | $2.64 \times 10^{-8}$ | $-0.07 \pm 0.01$ | 0.0059 [0.0025 to 0.0111] |
| Energy-adjusted Dietary Inflammatory Index | 5224 | 0.0087 | $0.03 \pm 0.01$ | 0.0013 [$8.88 \times 10^{-5}$ to 0.0041] |
| Dietary Inflammatory Index | 5224 | 0.9130 | $-0.0020 \pm 0.02$ | $2.31 \times 10^{-6}$ [$1.57 \times 10^{-7}$ to 0.0009] |
| EAT-Lancet Index: Vegetables | 5222 | 0.4630 | $-0.01 \pm 0.01$ | 0.0001 [$3.48 \times 10^{-7}$ to 0.0015] |
| EAT-Lancet Index: Fruits | 5222 | 0.0029 | $-0.04 \pm 0.01$ | 0.0017 [0.0002 to 0.0045] |
| EAT-Lancet Index: Unsaturated fats | 5222 | $1.82 \times 10^{-6}$ | $0.07 \pm 0.01$ | 0.0044 [0.0015 to 0.0086] |
| EAT-Lancet Index: Saturated fats | 5222 | $3.00 \times 10^{-6}$ | $-0.06 \pm 0.01$ | 0.0042 [0.0014 to 0.0084] |
| EAT-Lancet Index: Legumes (without soy) | 5222 | 0.9460 | $0.0016 \pm 0.02$ | $8.74 \times 10^{-7}$ [$1.79 \times 10^{-7}$ to 0.0010] |
| EAT-Lancet Index: Soy foods | 5222 | 0.4050 | $0.01 \pm 0.01$ | 0.0001 [$2.28 \times 10^{-7}$ to 0.0015] |
| EAT-Lancet Index: Nuts | 5222 | 0.0008 | $-0.05 \pm 0.01$ | 0.0021 [0.0003 to 0.0051] |
| EAT-Lancet Index: Whole grains | 5222 | $1.27 \times 10^{-5}$ | $-0.06 \pm 0.01$ | 0.0037 [0.0011 to 0.0079] |
| EAT-Lancet Index: Fish and shellfish | 5222 | 0.0001 | $0.06 \pm 0.02$ | 0.0028 [0.0006 to 0.0065] |
| EAT-Lancet Index: Beef and lamb | 5222 | $6.39 \times 10^{-16}$ | $-0.13 \pm 0.01$ | 0.0125 [0.0075 to 0.0193] |
| EAT-Lancet Index: Pork | 5222 | $1.86 \times 10^{-13}$ | $-0.10 \pm 0.01$ | 0.0103 [0.0056 to 0.0166] |
| EAT-Lancet Index: Chicken and other poultry | 5222 | 0.0120 | $-0.04 \pm 0.01$ | 0.0012 [$8.02 \times 10^{-5}$ to 0.0039] |
| EAT-Lancet Index: Eggs | 5222 | $1.33 \times 10^{-6}$ | $-0.07 \pm 0.01$ | 0.0045 [0.0016 to 0.0091] |
| EAT-Lancet Index: Dairy foods | 5222 | 0.1360 | $0.02 \pm 0.02$ | 0.0004 [$2.25 \times 10^{-6}$ to 0.0023] |
| EAT-Lancet Index: Potatoes and Tubers | 5222 | 0.6920 | $-0.01 \pm 0.01$ | $3.02 \times 10^{-5}$ [$1.38 \times 10^{-7}$ to 0.0012] |
| EAT-Lancet Index: Added sugars | 5222 | $4.59 \times 10^{-5}$ | $0.06 \pm 0.01$ | 0.0032 [0.0008 to 0.0067] |
| EAT-Lancet Index: Total score | 5222 | $2.74 \times 10^{-6}$ | $-0.07 \pm 0.01$ | 0.0042 [0.0014 to 0.0083] |

**Table S5.**
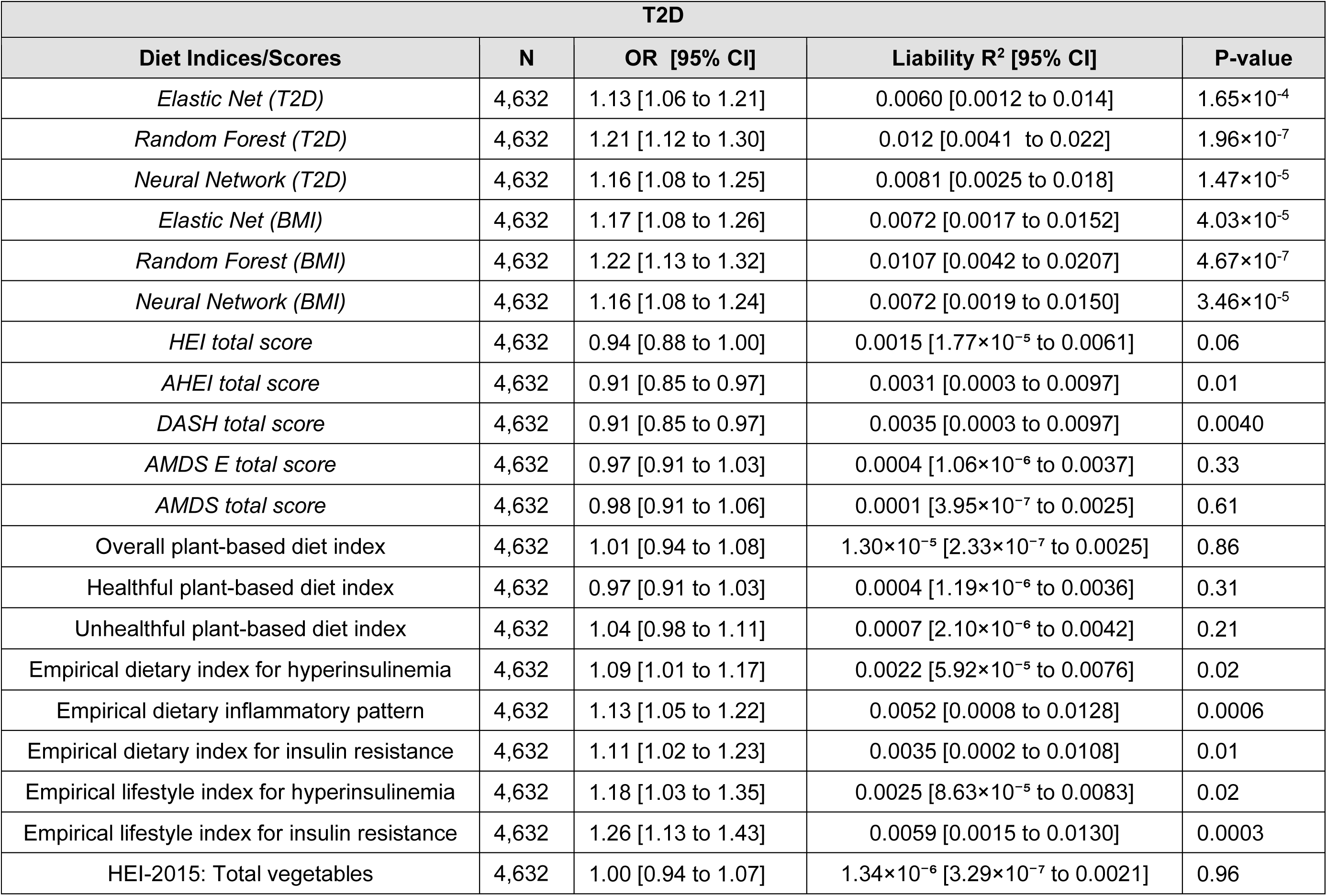
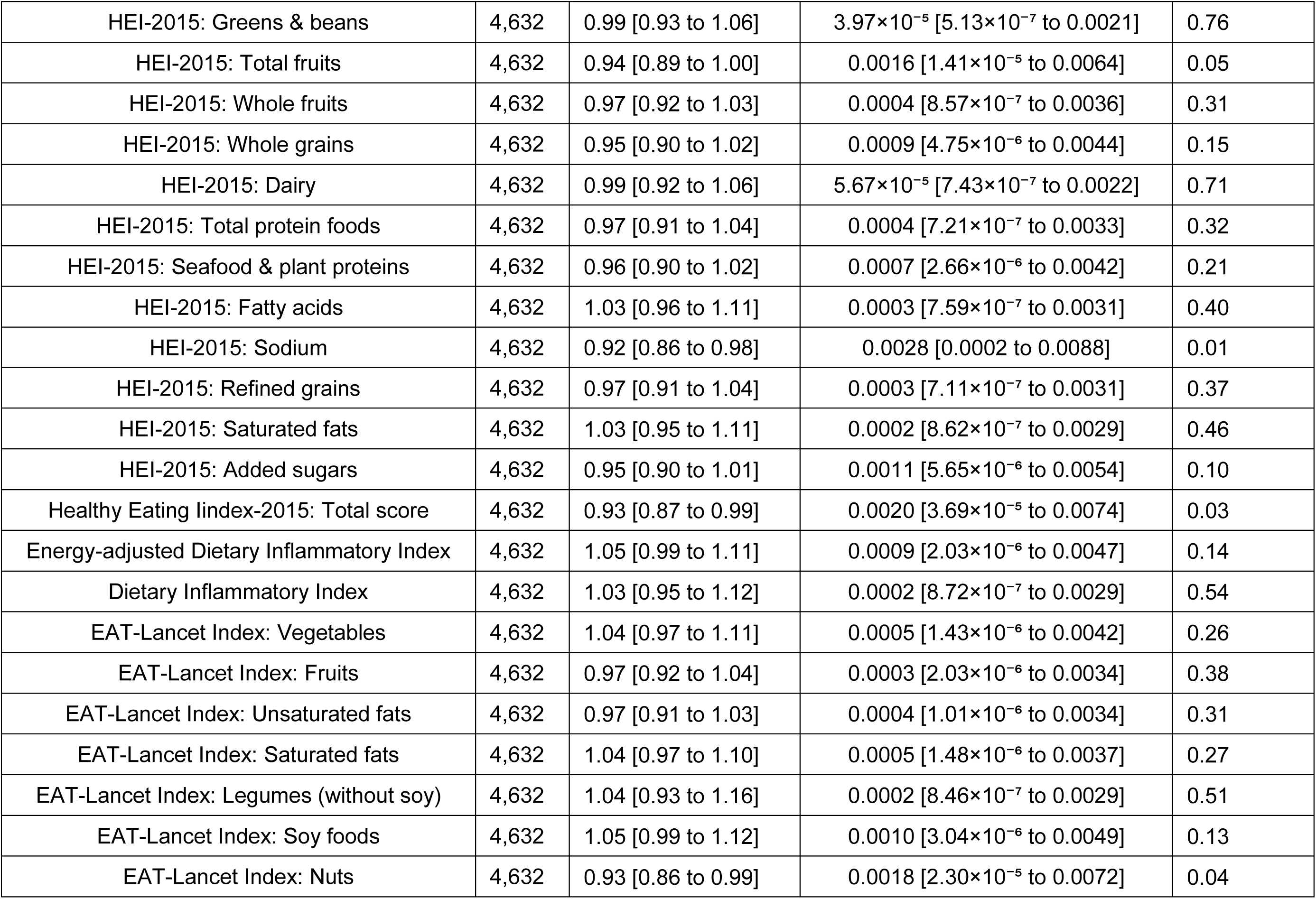
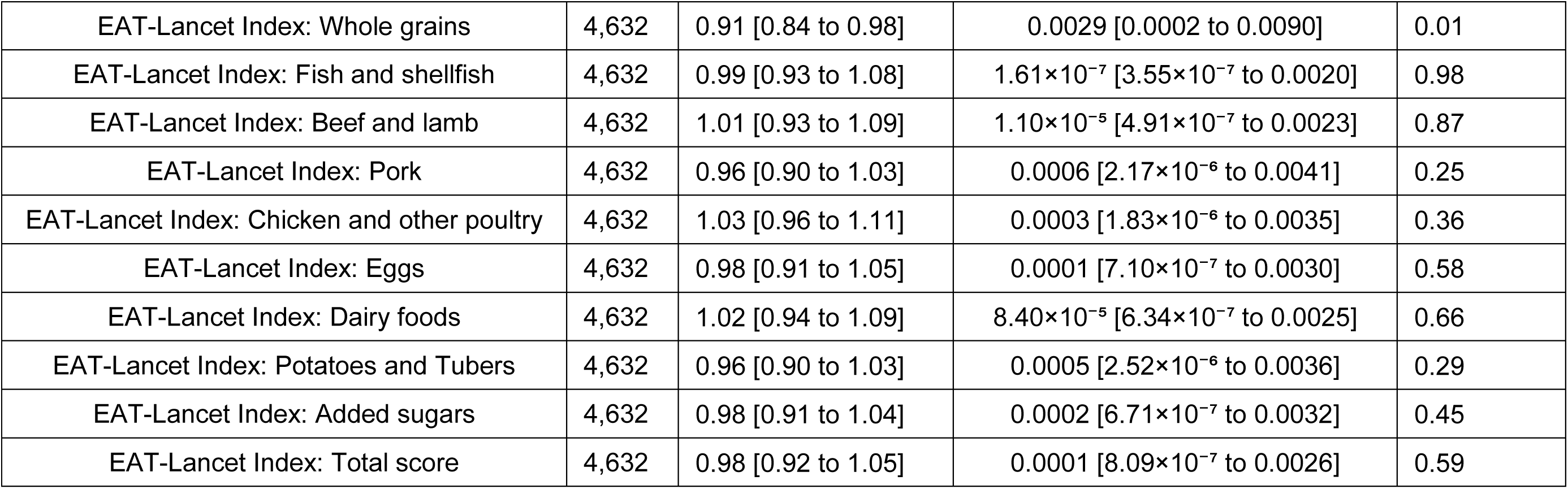
Association of dietary scores and indices to T2D in Native Hawaiians. Evaluations were conducted only in held-out Native Hawaiians that were not used to develop the diet score in this study. The associations between dietary scores/indices and T2D were adjusted for age, sex, BMI, and daily calorie intake.

**Table S6.**
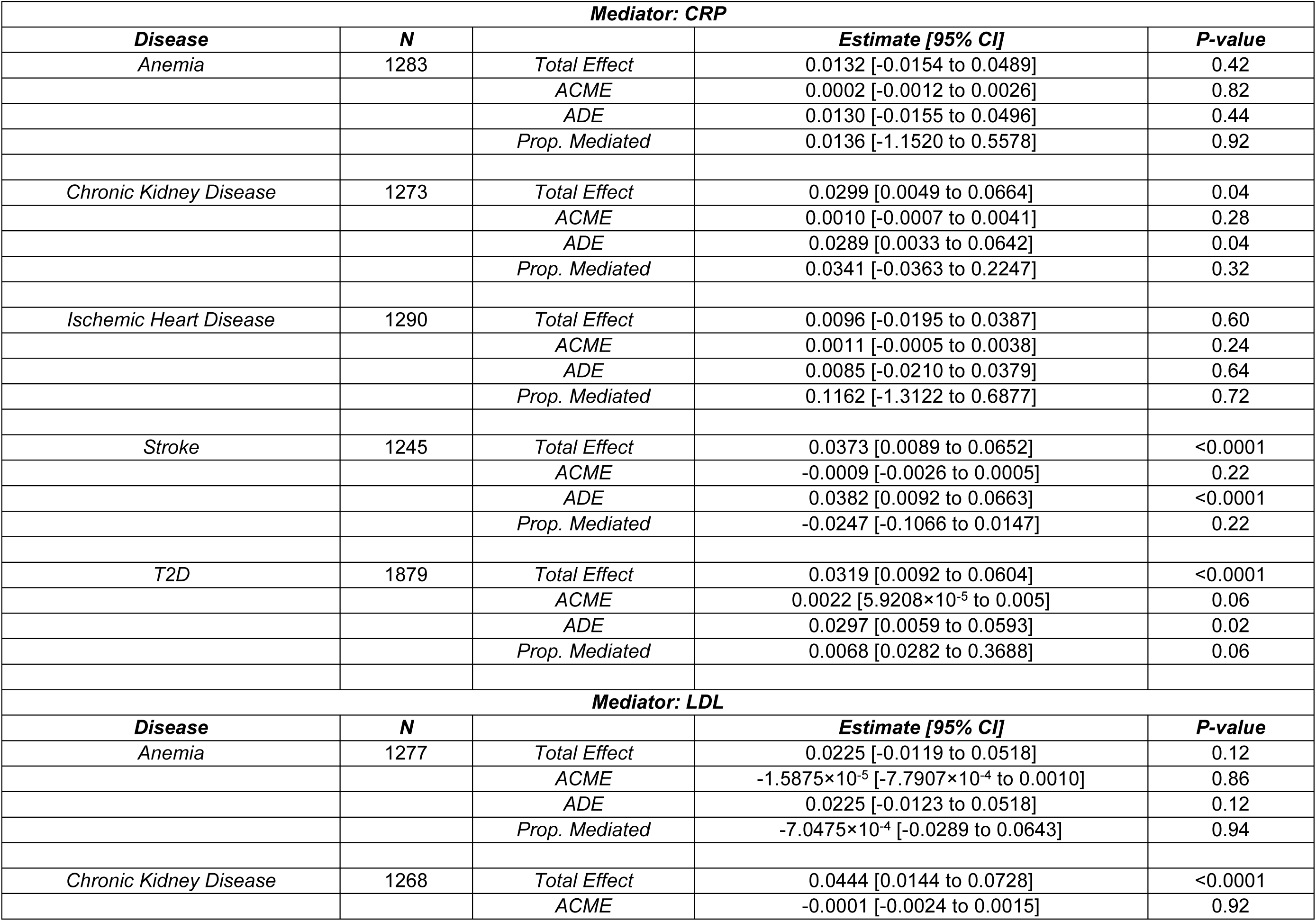
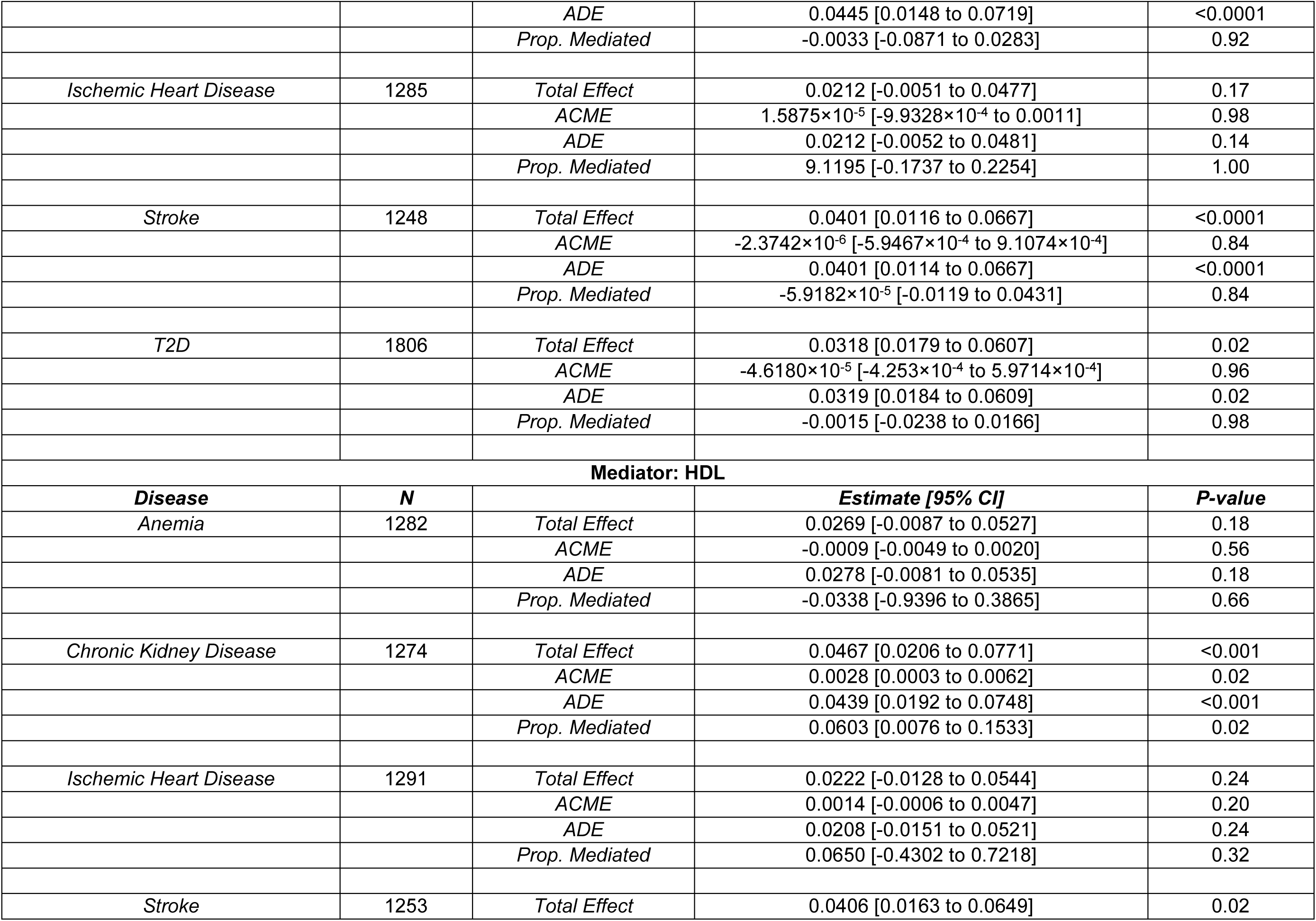
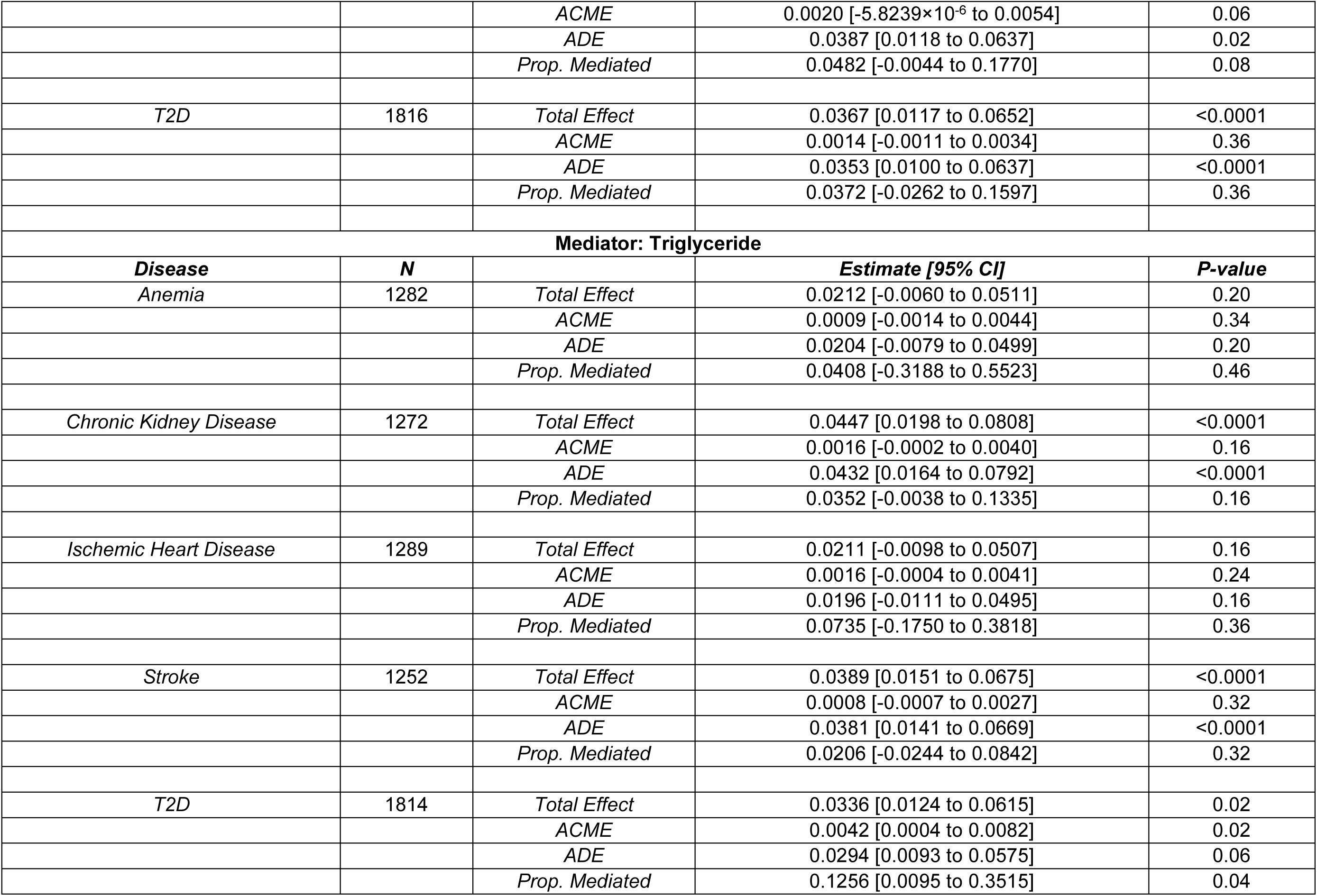
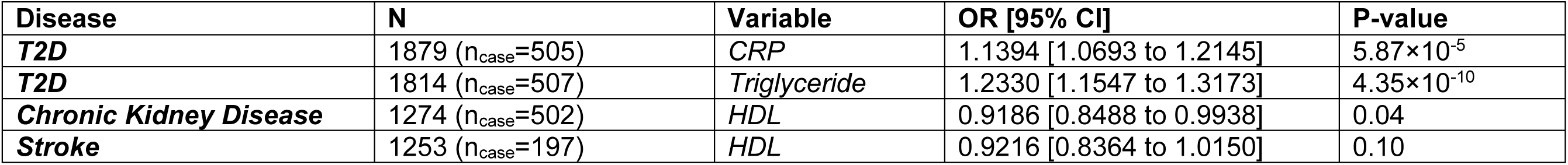
Potential mediators in Native Hawaiians. For the outcome phenotypes in which a significant association was observed between the BMI diet score and outcome, we tested the potential indirect effects of BMI diet score on disease phenotypes through mediation analysis with potential mediators. Specifically, we explored biomarkers that may reflect inflammation (C-reactive protein) and lipid pathways (low- and high-density lipoproteins, and triglycerides). The mediation analysis estimated total effect, which sums up direct and indirect effect of diet score, average causal mediated/indirect effect (ACME) of biomarkers, average direct effect (ADE) of diet score, and proportion of mediated effect. The marginal effects of mediator on phenotype were also reported as odds ratios after accounting for covariates (diet score, age, sex, BMI, daily energy intake (kcal/day), and first 10 PCs).

| **Mediator: CRP** |  |  |  |  |
| --- | --- | --- | --- | --- |
| **Disease** | **N** |  | **Estimate [95% CI]** | **P-value** |
| Anemia | 1283 | Total Effect | 0.0132 [-0.0154 to 0.0489] | 0.42 |
|  |  | ACME | 0.0002 [-0.0012 to 0.0026] | 0.82 |
|  |  | ADE | 0.0130 [-0.0155 to 0.0496] | 0.44 |
|  |  | Prop. Mediated | 0.0136 [-1.1520 to 0.5578] | 0.92 |
| Chronic Kidney Disease | 1273 | Total Effect | 0.0299 [0.0049 to 0.0664] | 0.04 |
|  |  | ACME | 0.0010 [-0.0007 to 0.0041] | 0.28 |
|  |  | ADE | 0.0289 [0.0033 to 0.0642] | 0.04 |
|  |  | Prop. Mediated | 0.0341 [-0.0363 to 0.2247] | 0.32 |
| Ischemic Heart Disease | 1290 | Total Effect | 0.0096 [-0.0195 to 0.0387] | 0.60 |
|  |  | ACME | 0.0011 [-0.0005 to 0.0038] | 0.24 |
|  |  | ADE | 0.0085 [-0.0210 to 0.0379] | 0.64 |
|  |  | Prop. Mediated | 0.1162 [-1.3122 to 0.6877] | 0.72 |
| Stroke | 1245 | Total Effect | 0.0373 [0.0089 to 0.0652] | <0.0001 |
|  |  | ACME | -0.0009 [-0.0026 to 0.0005] | 0.22 |
|  |  | ADE | 0.0382 [0.0092 to 0.0663] | <0.0001 |
|  |  | Prop. Mediated | -0.0247 [-0.1066 to 0.0147] | 0.22 |
| T2D | 1879 | Total Effect | 0.0319 [0.0092 to 0.0604] | <0.0001 |
|  |  | ACME | 0.0022 [5.9208×10<sup>-5</sup> to 0.005] | 0.06 |
|  |  | ADE | 0.0297 [0.0059 to 0.0593] | 0.02 |
|  |  | Prop. Mediated | 0.0068 [0.0282 to 0.3688] | 0.06 |
| **Mediator: LDL** |  |  |  |  |
| **Disease** | **N** |  | **Estimate [95% CI]** | **P-value** |
| Anemia | 1277 | Total Effect | 0.0225 [-0.0119 to 0.0518] | 0.12 |
|  |  | ACME | -1.5875×10<sup>-5</sup> [-7.7907×10<sup>-4</sup> to 0.0010] | 0.86 |
|  |  | ADE | 0.0225 [-0.0123 to 0.0518] | 0.12 |
|  |  | Prop. Mediated | -7.0475×10<sup>-4</sup> [-0.0289 to 0.0643] | 0.94 |
| Chronic Kidney Disease | 1268 | Total Effect | 0.0444 [0.0144 to 0.0728] | <0.0001 |
|  |  | ACME | -0.0001 [-0.0024 to 0.0015] | 0.92 |
|  |  | *ADE* | 0.0445 [0.0148 to 0.0719] | <0.0001 |
|  |  | *Prop. Mediated* | -0.0033 [-0.0871 to 0.0283] | 0.92 |
| *Ischemic Heart Disease* | 1285 | *Total Effect* | 0.0212 [-0.0051 to 0.0477] | 0.17 |
|  |  | *ACME* | 1.5875×10<sup>-5</sup> [-9.9328×10<sup>-4</sup> to 0.0011] | 0.98 |
|  |  | *ADE* | 0.0212 [-0.0052 to 0.0481] | 0.14 |
|  |  | *Prop. Mediated* | 9.1195 [-0.1737 to 0.2254] | 1.00 |
| *Stroke* | 1248 | *Total Effect* | 0.0401 [0.0116 to 0.0667] | <0.0001 |
|  |  | *ACME* | -2.3742×10<sup>-6</sup> [-5.9467×10<sup>-4</sup> to 9.1074×10<sup>-4</sup>] | 0.84 |
|  |  | *ADE* | 0.0401 [0.0114 to 0.0667] | <0.0001 |
|  |  | *Prop. Mediated* | -5.9182×10<sup>-5</sup> [-0.0119 to 0.0431] | 0.84 |
| *T2D* | 1806 | *Total Effect* | 0.0318 [0.0179 to 0.0607] | 0.02 |
|  |  | *ACME* | -4.6180×10<sup>-5</sup> [-4.253×10<sup>-4</sup> to 5.9714×10<sup>-4</sup>] | 0.96 |
|  |  | *ADE* | 0.0319 [0.0184 to 0.0609] | 0.02 |
|  |  | *Prop. Mediated* | -0.0015 [-0.0238 to 0.0166] | 0.98 |
| **Mediator: HDL** |  |  |  |  |
| ***Disease*** | ***N*** |  | ***Estimate [95% CI]*** | ***P-value*** |
| *Anemia* | 1282 | *Total Effect* | 0.0269 [-0.0087 to 0.0527] | 0.18 |
|  |  | *ACME* | -0.0009 [-0.0049 to 0.0020] | 0.56 |
|  |  | *ADE* | 0.0278 [-0.0081 to 0.0535] | 0.18 |
|  |  | *Prop. Mediated* | -0.0338 [-0.9396 to 0.3865] | 0.66 |
| *Chronic Kidney Disease* | 1274 | *Total Effect* | 0.0467 [0.0206 to 0.0771] | <0.001 |
|  |  | *ACME* | 0.0028 [0.0003 to 0.0062] | 0.02 |
|  |  | *ADE* | 0.0439 [0.0192 to 0.0748] | <0.001 |
|  |  | *Prop. Mediated* | 0.0603 [0.0076 to 0.1533] | 0.02 |
| *Ischemic Heart Disease* | 1291 | *Total Effect* | 0.0222 [-0.0128 to 0.0544] | 0.24 |
|  |  | *ACME* | 0.0014 [-0.0006 to 0.0047] | 0.20 |
|  |  | *ADE* | 0.0208 [-0.0151 to 0.0521] | 0.24 |
|  |  | *Prop. Mediated* | 0.0650 [-0.4302 to 0.7218] | 0.32 |
| *Stroke* | 1253 | *Total Effect* | 0.0406 [0.0163 to 0.0649] | 0.02 |
|  |  | ACME | 0.0020 [-5.8239×10<sup>-6</sup> to 0.0054] | 0.06 |
|  |  | ADE | 0.0387 [0.0118 to 0.0637] | 0.02 |
|  |  | Prop. Mediated | 0.0482 [-0.0044 to 0.1770] | 0.08 |
| T2D | 1816 | Total Effect | 0.0367 [0.0117 to 0.0652] | <0.0001 |
|  |  | ACME | 0.0014 [-0.0011 to 0.0034] | 0.36 |
|  |  | ADE | 0.0353 [0.0100 to 0.0637] | <0.0001 |
|  |  | Prop. Mediated | 0.0372 [-0.0262 to 0.1597] | 0.36 |
| Mediator: Triglyceride |  |  |  |  |
| **Disease** | **N** |  | **Estimate [95% CI]** | **P-value** |
| Anemia | 1282 | Total Effect | 0.0212 [-0.0060 to 0.0511] | 0.20 |
|  |  | ACME | 0.0009 [-0.0014 to 0.0044] | 0.34 |
|  |  | ADE | 0.0204 [-0.0079 to 0.0499] | 0.20 |
|  |  | Prop. Mediated | 0.0408 [-0.3188 to 0.5523] | 0.46 |
| Chronic Kidney Disease | 1272 | Total Effect | 0.0447 [0.0198 to 0.0808] | <0.0001 |
|  |  | ACME | 0.0016 [-0.0002 to 0.0040] | 0.16 |
|  |  | ADE | 0.0432 [0.0164 to 0.0792] | <0.0001 |
|  |  | Prop. Mediated | 0.0352 [-0.0038 to 0.1335] | 0.16 |
| Ischemic Heart Disease | 1289 | Total Effect | 0.0211 [-0.0098 to 0.0507] | 0.16 |
|  |  | ACME | 0.0016 [-0.0004 to 0.0041] | 0.24 |
|  |  | ADE | 0.0196 [-0.0111 to 0.0495] | 0.16 |
|  |  | Prop. Mediated | 0.0735 [-0.1750 to 0.3818] | 0.36 |
| Stroke | 1252 | Total Effect | 0.0389 [0.0151 to 0.0675] | <0.0001 |
|  |  | ACME | 0.0008 [-0.0007 to 0.0027] | 0.32 |
|  |  | ADE | 0.0381 [0.0141 to 0.0669] | <0.0001 |
|  |  | Prop. Mediated | 0.0206 [-0.0244 to 0.0842] | 0.32 |
| T2D | 1814 | Total Effect | 0.0336 [0.0124 to 0.0615] | 0.02 |
|  |  | ACME | 0.0042 [0.0004 to 0.0082] | 0.02 |
|  |  | ADE | 0.0294 [0.0093 to 0.0575] | 0.06 |
|  |  | Prop. Mediated | 0.1256 [0.0095 to 0.3515] | 0.04 |

Table S6. Potential mediators in Native Hawaiians.
| **Disease** | **N** | **Variable** | **OR [95% CI]** | **P-value** |
| --- | --- | --- | --- | --- |
| ***T2D*** | 1879 (n<sub>case</sub>=505) | *CRP* | 1.1394 [1.0693 to 1.2145] | 5.87×10<sup>-5</sup> |
| ***T2D*** | 1814 (n<sub>case</sub>=507) | *Triglyceride* | 1.2330 [1.1547 to 1.3173] | 4.35×10<sup>-10</sup> |
| ***Chronic Kidney Disease*** | 1274 (n<sub>case</sub>=502) | *HDL* | 0.9186 [0.8488 to 0.9938] | 0.04 |
| ***Stroke*** | 1253 (n<sub>case</sub>=197) | *HDL* | 0.9216 [0.8364 to 1.0150] | 0.10 |

**Table S7.**
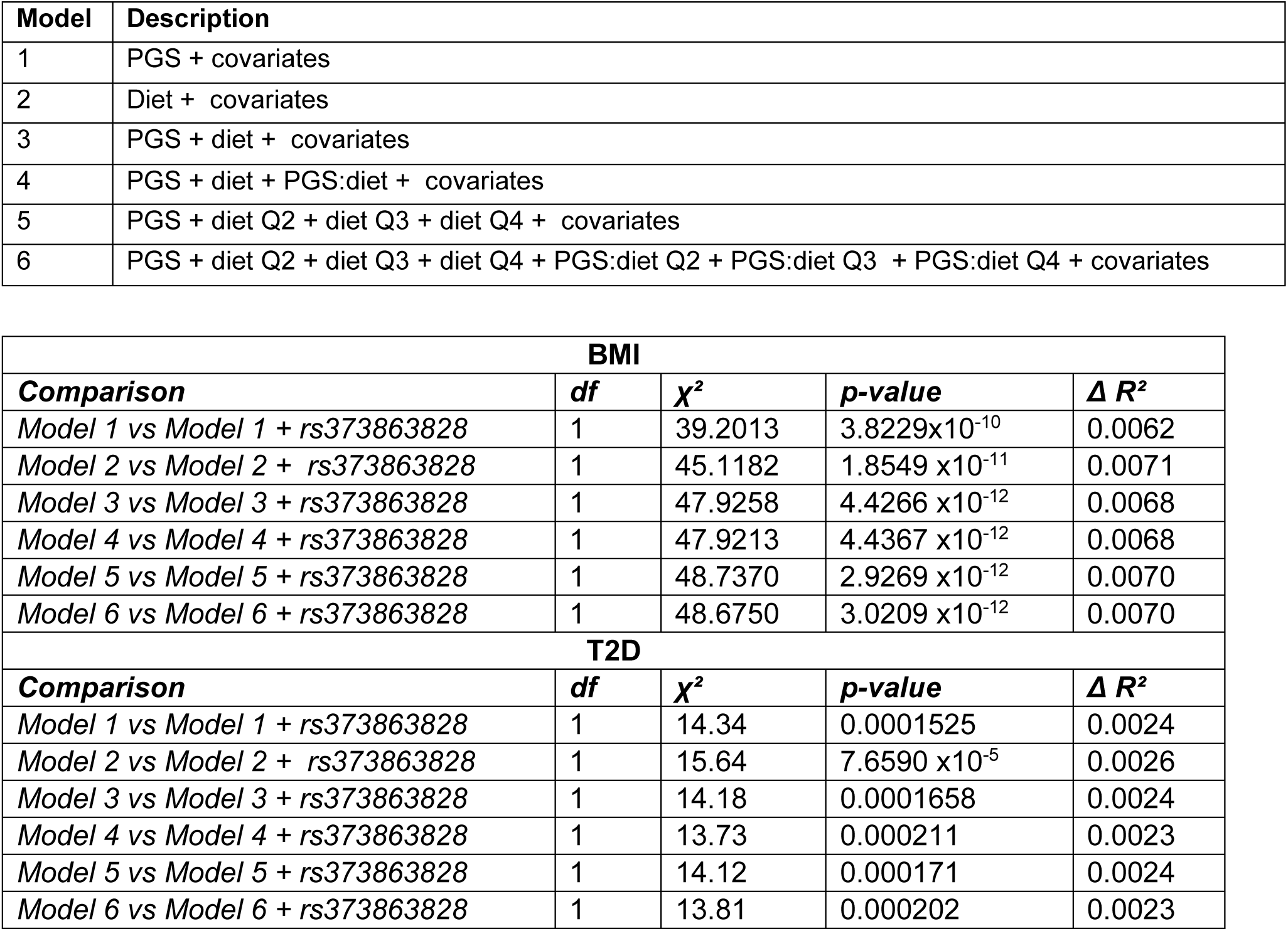
Inclusion of *CREBRF* variant in risk prediction models for BMI and T2D. Performance of model with and without a population-specific variant, *rs373863828,* was compared using the log-likelihood ratio test. * All models for T2D were adjusted for age, sex, BMI, smoking status, physical activity, education level, daily energy intake (kcal/day), and first 10 PCs. All BMI models included smoking status, physical activity, education level, daily energy intake (kcal/day) and first 10 PCs.

| Model | Description |
| --- | --- |
| 1 | PGS + covariates |
| 2 | Diet + covariates |
| 3 | PGS + diet + covariates |
| 4 | PGS + diet + PGS:diet + covariates |
| 5 | PGS + diet Q2 + diet Q3 + diet Q4 + covariates |
| 6 | PGS + diet Q2 + diet Q3 + diet Q4 + PGS:diet Q2 + PGS:diet Q3 + PGS:diet Q4 + covariates |

Table S7. Inclusion of *CREBRF* variant in risk prediction models for BMI and T2D.
| BMI |  |  |  |  |
| --- | --- | --- | --- | --- |
| Comparison | df | $\chi^2$ | p-value | $\Delta R^2$ |
| Model 1 vs Model 1 + rs373863828 | 1 | 39.2013 | 3.8229x10<sup>-10</sup> | 0.0062 |
| Model 2 vs Model 2 + rs373863828 | 1 | 45.1182 | 1.8549 x10<sup>-11</sup> | 0.0071 |
| Model 3 vs Model 3 + rs373863828 | 1 | 47.9258 | 4.4266 x10<sup>-12</sup> | 0.0068 |
| Model 4 vs Model 4 + rs373863828 | 1 | 47.9213 | 4.4367 x10<sup>-12</sup> | 0.0068 |
| Model 5 vs Model 5 + rs373863828 | 1 | 48.7370 | 2.9269 x10<sup>-12</sup> | 0.0070 |
| Model 6 vs Model 6 + rs373863828 | 1 | 48.6750 | 3.0209 x10<sup>-12</sup> | 0.0070 |
| T2D |  |  |  |  |
| Comparison | df | $\chi^2$ | p-value | $\Delta R^2$ |
| Model 1 vs Model 1 + rs373863828 | 1 | 14.34 | 0.0001525 | 0.0024 |
| Model 2 vs Model 2 + rs373863828 | 1 | 15.64 | 7.6590 x10<sup>-5</sup> | 0.0026 |
| Model 3 vs Model 3 + rs373863828 | 1 | 14.18 | 0.0001658 | 0.0024 |
| Model 4 vs Model 4 + rs373863828 | 1 | 13.73 | 0.000211 | 0.0023 |
| Model 5 vs Model 5 + rs373863828 | 1 | 14.12 | 0.000171 | 0.0024 |
| Model 6 vs Model 6 + rs373863828 | 1 | 13.81 | 0.000202 | 0.0023 |

## REFERENCES

1. Alberti KGMM, Zimmet P, Shaw J. (2005) The metabolic syndrome—a new worldwide definition. The Lancet 366(9491):1059–1062. doi: 10.1016/S0140-6736(05)67402-8

2. Beltrán-Sánchez H, Harhay MO, Harhay MM, McElligott S. Prevalence and trends of metabolic syndrome in the adult U.S. population, 1999-2010. J Am Coll Cardiol. 2013 Aug 20;62(8):697–703. doi: 10.1016/j.jacc.2013.05.064. Epub 2013 Jun 27. PMID: 23810877; PMCID: PMC3756561.

3. American Heart Association (2024). What is Metabolic Syndrome? Retrieved from https://www.heart.org/en/health-topics/metabolic-syndrome/about-metabolic-syndrome

4. Prasad GV. Metabolic syndrome and chronic kidney disease: Current status and future directions. World J Nephrol. 2014 Nov 6;3(4):210–9. doi: 10.5527/wjn.v3.i4.210. PMID: 25374814; PMCID: PMC4220353.

5. Li W, Chen D, Peng Y, Lu Z, Kwan MP, Tse LA. Association Between Metabolic Syndrome and Mortality: Prospective Cohort Study. JMIR Public Health Surveill. 2023 Sep 5;9:e44073. doi: 10.2196/44073. PMID: 37669100; PMCID: PMC10509744.

6. World Health Organization (2025). Obesity and overweight. Fact Sheet.

7. 7. CDC (2024). National Diabetes Statistics Report. Centers for Disease Control and Prevention

8. CDC (2025). Diabetes in Young People Is on the Rise. Centers for Disease Control and Prevention.

9. Yengo, L., Liang, Y., 23 and Me Research Team, Wang, X., Granka, J. M., Evans, D. M., Sidorenko, J., & Visscher, P. M. (2025). Within-family heritability estimates for behavioural and disease phenotypes from 500,000 sibling pairs of diverse ancestries. medRxiv. 10.1101/2025.09.17.25336022

10. Xiang, R., Kelemen, M., Xu, Y. et al. Recent advances in polygenic scores: translation, equitability, methods and FAIR tools. Genome Med 16, 33 (2024). 10.1186/s13073-024-01304-9

11. Truong, B. et al. (2024). Integrative polygenic risk score improves the prediction accuracy of complex traits and diseases. Cell Genomics, 4(3), 100504

12. Slunecka, J.L., van der Zee, M.D., Beck, J.J. et al. Implementation and implications for polygenic risk scores in healthcare. Hum Genomics 15, 46 (2021). 10.1186/s40246-021-00339-y

13. Duncan, L., Shen, H., Gelaye, B. et al. Analysis of polygenic risk score usage and performance in diverse human populations. Nat Commun 10, 3328 (2019). 10.1038/s41467-019-11112-0

14. Wang, Y., Guo, J., Ni, G. et al. Theoretical and empirical quantification of the accuracy of polygenic scores in ancestry divergent populations. Nat Commun 11, 3865 (2020). 10.1038/s41467-020-17719-y

15. Lo Y-C, Tian H, Chan TF, Jeon S, Alatorre K, Dinh BL, Maskarinec G, Taparra K, Nakatsuka N, Yu M, Chen C-Y, Lin Y-F, Wilkens LR, Le Marchand L, Haiman CA, Chiang CWK. (2025) The accuracy of polygenic score models for BMI and Type II diabetes in the Native Hawaiian population. Communications Biology 8(1):651. doi: 10.1038/s42003-025-08050-7

16. Jeon S, Lo YC, Morimoto LM, Metayer C, Ma X, Wiemels JL, de Smith AJ, Chiang CWK. Evaluating genomic polygenic risk scores for childhood acute lymphoblastic leukemia in Latinos. HGG Adv. 2023 Oct 12;4(4):100239. doi: 10.1016/j.xhgg.2023.100239. Epub 2023 Sep 14. PMID: 37710962; PMCID: PMC10550840.

17. Ambroselli D, Masciulli F, Romano E, Catanzaro G, Besharat ZM, Massari MC, Ferretti E, Migliaccio S, Izzo L, Ritieni A, Grosso M, Formichi C, Dotta F, Frigerio F, Barbiera E, Giusti AM, Ingallina C, Mannina L. New Advances in Metabolic Syndrome, from Prevention to Treatment: The Role of Diet and Food. Nutrients. 2023 Jan 26;15(3):640. doi: 10.3390/nu15030640. PMID: 36771347; PMCID: PMC9921449.

18. Clemente-Suárez VJ, Beltrán-Velasco AI, Redondo-Flórez L, Martín-Rodríguez A, Tornero-Aguilera JF. Global Impacts of Western Diet and Its Effects on Metabolism and Health: A Narrative Review. Nutrients. 2023 Jun 14;15(12):2749. doi: 10.3390/nu15122749. PMID: 37375654; PMCID: PMC10302286.

19. Castro-Barquero S, Ruiz-León AM, Sierra-Pérez M, Estruch R, Casas R. Dietary Strategies for Metabolic Syndrome: A Comprehensive Review. Nutrients. 2020 Sep 29;12(10):2983. doi: 10.3390/nu12102983. PMID: 33003472; PMCID: PMC7600579.

20. Kastorini CM, Milionis HJ, Esposito K, Giugliano D, Goudevenos JA, Panagiotakos DB. The effect of Mediterranean diet on metabolic syndrome and its components: a meta-analysis of 50 studies and 534,906 individuals. J Am Coll Cardiol. 2011 Mar 15;57(11):1299–313. doi: 10.1016/j.jacc.2010.09.073. PMID: 21392646.

21. Klapp R, Laxamana JA, Shvetsov YB, Park SY, Kanehara R, Setiawan VW, Danquah I, Le Marchand L, Maskarinec G. The EAT-Lancet Diet Index Is Associated with Lower Obesity and Incidence of Type 2 Diabetes in the Multiethnic Cohort. J Nutr. 2024 Nov;154(11):3407–3415. doi: 10.1016/j.tjnut.2024.06.018. Epub 2024 Jul 15. PMID: 39019161; PMCID: PMC11600087.

22. Park SY, Boushey CJ, Wilkens LR, Haiman CA, Le Marchand L. High-Quality Diets Associate With Reduced Risk of Colorectal Cancer: Analyses of Diet Quality Indexes in the Multiethnic Cohort. Gastroenterology. 2017 Aug;153(2):386–394.e2. doi: 10.1053/j.gastro.2017.04.004. Epub 2017 Apr 17. PMID: 28428143; PMCID: PMC5526717.

23. CDC/HHS Office of Minority Health data. https://minorityhealth.hhs.gov/diabetes-and-native-hawaiianspacific-islanders

24. Karter AJ, Schillinger D, Adams AS, Moffet HH, Liu J, Adler NE, Kanaya AM. Elevated rates of diabetes in Pacific Islanders and Asian subgroups: The Diabetes Study of Northern California (DISTANCE). Diabetes Care. 2013 Mar;36(3):574–9. doi: 10.2337/dc12-0722. Epub 2012 Oct 15. PMID: 23069837; PMCID: PMC3579366.

25. Kolonel LN, Henderson BE, Hankin JH, Nomura AM, Wilkens LR, Pike MC, Stram DO, Monroe KR, Earle ME, Nagamine FS. A multiethnic cohort in Hawaii and Los Angeles: baseline characteristics. Am J Epidemiol. 2000 Feb 15;151(4):346–57. doi: 10.1093/oxfordjournals.aje.a010213. PMID: 10695593; PMCID: PMC4482109.

26. Stram DO, Hankin JH, Wilkens LR, Pike MC, Monroe KR, Park S, Henderson BE, Nomura AM, Earle ME, Nagamine FS, Kolonel LN. Calibration of the dietary questionnaire for a multiethnic cohort in Hawaii and Los Angeles. Am J Epidemiol. 2000 Feb 15;151(4):358–70. doi: 10.1093/oxfordjournals.aje.a010214. PMID: 10695594; PMCID: PMC4482461.

27. Bogumil D, Sheng X, Wan P, Xia L, Pooler L, Cheng I, Streicher S, Huang BZ, Chen F, Stram D, Shen S, King G, Chiang CWK, Ongaco C, Adams M, McMullen I, Zhang P, Ling H, Mawhinney M, Doheny KF, Marchand LL, Wilkens LR, Haiman CA, Conti DV. The Multiethnic Cohort: A Resource for the study of Genetic and non-Genetic Cancer Risk Across Populations. medRxiv [Preprint]. 2025 Jun 11:2025.06.09.25328993. doi: 10.1101/2025.06.09.25328993. PMID: 40585109; PMCID: PMC12204407.

28. Sheng X, Xia L, Cahoon JL, Conti DV, Haiman CA, Kachuri L, Chiang CWK. Inverted genomic regions between reference genome builds in humans impact imputation accuracy and decrease the power of association testing. HGG Adv. 2022 Nov 11;4(1):100159. doi: 10.1016/j.xhgg.2022.100159. PMID: 36465187; PMCID: PMC9709082.

29. Loh, P.R., Danecek, P., Palamara, P.F., Fuchsberger, C., Reshef, Y.A., Finucane, H.K., Schoenherr, S., Forer, L., McCarthy, S., Abecasis, G.R., Durbin, R., & Price, A.L. (2016). Reference-based phasing using the Haplotype Reference Consortium panel. Nature Genetics, 48(11), 1443–1448. 10.1038/ng.3679

30. Dinh, B.L., Tang, E., Taparra, K., Nakatsuka, N., Chen, F., & Chiang, C.W.K. (2024). Recombination map tailored to Native Hawaiians may improve robustness of genomic scans for positive selection. Human Genetics, 143(1), 85–99. 10.1007/s00439-023-02625-2

31. Taliun, D., Harris, D.N., Kessler, M.D., Carlson, J., Szpiech, Z.A., Torres, R., Taliun, S.A.G., Corvelo, A., Gogarten, S.M., Kang, H.M., et al. (2021). Sequencing of 53,831 diverse genomes from the NHLBI TOPMed Program. Nature, 589(7845), 290–299. 10.1038/s41586-021-03205-y

32. Manichaikul A, Mychaleckyj JC, Rich SS, Daly K, Sale M, Chen WM (2010) Robust relationship inference in genome-wide association studies. Bioinformatics 26(22):2867–2873

33. Conomos MP, Miller MB, Thornton TA. Robust inference of population structure for ancestry prediction and correction of stratification in the presence of relatedness. Genet Epidemiol. 2015 May;39(4):276–93. doi: 10.1002/gepi.21896. Epub 2015 Mar 23. PMID: 25810074; PMCID: PMC4836868.

34. Conomos MP, Reiner AP, Weir BS, Thornton TA. Model-free Estimation of Recent Genetic Relatedness. Am J Hum Genet. 2016 Jan 7;98(1):127–48. doi: 10.1016/j.ajhg.2015.11.022. PMID: 26748516; PMCID: PMC4716688.

35. Stephanie M Gogarten, Tamar Sofer, Han Chen, Chaoyu Yu, Jennifer A Brody, Timothy A Thornton, Kenneth M Rice, Matthew P Conomos, Genetic association testing using the GENESIS R/Bioconductor package, *Bioinformatics*, Volume 35, Issue 24, December 2019, Pages 5346–5348, 10.1093/bioinformatics/btz567

36. Y Ruan, YF Lin, YCA Feng, CY Chen, M Lam, Z Guo, Stanley Global Asia Initiatives, L He, A Sawa, AR Martin, S Qin, H Huang, T Ge. Improving polygenic prediction in ancestrally diverse populations. Nature Genetics, 54:573–580, 2022.

37. Hoggart, C.J., Choi, S.W., García-González, J. et al. BridgePRS leverages shared genetic effects across ancestries to increase polygenic risk score portability. Nat Genet 56, 180–186 (2024). 10.1038/s41588-023-01583-9

38. Suzuki K, Hatzikotoulas K, Southam L, Taylor HJ, Yin X, Lorenz KM, Mandla R, Huerta-Chagoya A, Melloni GEM, Kanoni S, Rayner NW, Bocher O, Arruda AL, Sonehara K, Namba S, Lee SSK, Preuss MH, Petty LE, Schroeder P, Vanderwerff B, Kals M, Bragg F, Lin K, Guo X, Zhang W, Yao J, Kim YJ, Graff M, Takeuchi F, Nano J, Lamri A, Nakatochi M, Moon S, Scott RA, Cook JP, Lee JJ, Pan I, Taliun D, Parra EJ, Chai JF, Bielak LF, Tabara Y, Hai Y, Thorleifsson G, Grarup N, Sofer T, Wuttke M, Sarnowski C, Gieger C, Nousome D, Trompet S, Kwak SH, Long J, Sun M, Tong L, Chen WM, Nongmaithem SS, Noordam R, Lim VJY, Tam CHT, Joo YY, Chen CH, Raffield LM, Prins BP, Nicolas A, Yanek LR, Chen G, Brody JA, Kabagambe E, An P, Xiang AH, Choi HS, Cade BE, Tan J, Broadaway KA, Williamson A, Kamali Z, Cui J, Thangam M, Adair LS, Adeyemo A, Aguilar-Salinas CA, Ahluwalia TS, Anand SS, Bertoni A, Bork-Jensen J, Brandslund I, Buchanan TA, Burant CF, Butterworth AS, Canouil M, Chan JCN, Chang LC, Chee ML, Chen J, Chen SH, Chen YT, Chen Z, Chuang LM, Cushman M, Danesh J, Das SK, de Silva HJ, Dedoussis G, Dimitrov L, Doumatey AP, Du S, Duan Q, Eckardt KU, Emery LS, Evans DS, Evans MK, Fischer K, Floyd JS, Ford I, Franco OH, Frayling TM, Freedman BI, Genter P, Gerstein HC, Giedraitis V, González-Villalpando C, González-Villalpando ME, Gordon-Larsen P, Gross M, Guare LA, Hackinger S, Hakaste L, Han S, Hattersley AT, Herder C, Horikoshi M, Howard AG, Hsueh W, Huang M, Huang W, Hung YJ, Hwang MY, Hwu CM, Ichihara S, Ikram MA, Ingelsson M, Islam MT, Isono M, Jang HM, Jasmine F, Jiang G, Jonas JB, Jørgensen T, Kamanu FK, Kandeel FR, Kasturiratne A, Katsuya T, Kaur V, Kawaguchi T, Keaton JM, Kho AN, Khor CC, Kibriya MG, Kim DH, Kronenberg F, Kuusisto J, Läll K, Lange LA, Lee KM, Lee MS, Lee NR, Leong A, Li L, Li Y, Li-Gao R, Ligthart S, Lindgren CM, Linneberg A, Liu CT, Liu J, Locke AE, Louie T, Luan J, Luk AO, Luo X, Lv J, Lynch JA, Lyssenko V, Maeda S, Mamakou V, Mansuri SR, Matsuda K, Meitinger T, Melander O, Metspalu A, Mo H, Morris AD, Moura FA, Nadler JL, Nalls MA, Nayak U, Ntalla I, Okada Y, Orozco L, Patel SR, Patil S, Pei P, Pereira MA, Peters A, Pirie FJ, Polikowsky HG, Porneala B, Prasad G, Rasmussen-Torvik LJ, Reiner AP, Roden M, Rohde R, Roll K, Sabanayagam C, Sandow K, Sankareswaran A, Sattar N, Schönherr S, Shahriar M, Shen B, Shi J, Shin DM, Shojima N, Smith JA, So WY, Stančáková A, Steinthorsdottir V, Stilp AM, Strauch K, Taylor KD, Thorand B, Thorsteinsdottir U, Tomlinson B, Tran TC, Tsai FJ, Tuomilehto J, Tusie-Luna T, Udler MS, Valladares-Salgado A, van Dam RM, van Klinken JB, Varma R, Wacher-Rodarte N, Wheeler E, Wickremasinghe AR, van Dijk KW, Witte DR, Yajnik CS, Yamamoto K, Yamamoto K, Yoon K, Yu C, Yuan JM, Yusuf S, Zawistowski M, Zhang L, Zheng W; VA Million Veteran Program; Raffel LJ, Igase M, Ipp E, Redline S, Cho YS, Lind L, Province MA, Fornage M, Hanis CL, Ingelsson E, Zonderman AB, Psaty BM, Wang YX, Rotimi CN, Becker DM, Matsuda F, Liu Y, Yokota M, Kardia SLR, Peyser PA, Pankow JS, Engert JC, Bonnefond A, Froguel P, Wilson JG, Sheu WHH, Wu JY, Hayes MG, Ma RCW, Wong TY, Mook-Kanamori DO, Tuomi T, Chandak GR, Collins FS, Bharadwaj D, Paré G, Sale MM, Ahsan H, Motala AA, Shu XO, Park KS, Jukema JW, Cruz M, Chen YI, Rich SS, McKean-Cowdin R, Grallert H, Cheng CY, Ghanbari M, Tai ES, Dupuis J, Kato N, Laakso M, Köttgen A, Koh WP, Bowden DW, Palmer CNA, Kooner JS, Kooperberg C, Liu S, North KE, Saleheen D, Hansen T, Pedersen O, Wareham NJ, Lee J, Kim BJ, Millwood IY, Walters RG, Stefansson K, Ahlqvist E, Goodarzi MO, Mohlke KL, Langenberg C, Haiman CA, Loos RJF, Florez JC, Rader DJ, Ritchie MD, Zöllner S, Mägi R, Marston NA, Ruff CT, van Heel DA, Finer S, Denny JC, Yamauchi T, Kadowaki T, Chambers JC, Ng MCY, Sim X, Below JE, Tsao PS, Chang KM, McCarthy MI, Meigs JB, Mahajan A, Spracklen CN, Mercader JM, Boehnke M, Rotter JI, Vujkovic M, Voight BF, Morris AP, Zeggini E. Genetic drivers of heterogeneity in type 2 diabetes pathophysiology. Nature. 2024 Mar;627(8003):347–357. doi: 10.1038/s41586-024-07019-6. Epub 2024 Feb 19. PMID: 38374256; PMCID: PMC10937372.

39. Lee SH, Goddard ME, Wray NR, Visscher PM. A better coefficient of determination for genetic profile analysis. Genet Epidemiol. 2012 Apr;36(3):214–24. doi: 10.1002/gepi.21614. PMID: 22714935.

40. Berisa T, Pickrell JK. (2016) Approximately independent linkage disequilibrium blocks in human populations. Bioinformatics 32(2):283–285. doi: 10.1093/bioinformatics/btv546

41. Liese AD, Krebs-Smith SM, Subar AF, George SM, Harmon BE, Neuhouser ML, Boushey CJ, Schap TE, Reedy J. The Dietary Patterns Methods Project: synthesis of findings across cohorts and relevance to dietary guidance. J Nutr. 2015 Mar;145(3):393–402. doi: 10.3945/jn.114.205336. Epub 2015 Jan 21. PMID: 25733454; PMCID: PMC4336525.

42. Castro JL, Mantas CJ, Benítez JM. Neural networks with a continuous squashing function in the output are universal approximators. Neural Netw. 2000 Jul;13(6):561–3. doi: 10.1016/s0893-6080(00)00031-9. PMID: 10987509.

43. arXiv:1603.04467v2

44. Setiawan VW, Virnig BA, Porcel J, Henderson BE, Le Marchand L, Wilkens LR, et al. Linking data from the Multiethnic Cohort Study to Medicare data: linkage results and application to chronic disease research. Am J Epidemiol. 2015;181: 917–919. pmid: 25841869

45. Wojcik GL, Graff M, Nishimura KK, Tao R, Haessler J, Gignoux CR, Highland HM, Patel YM, Sorokin EP, Avery CL, Belbin GM, Bien SA, Cheng I, Cullina S, Hodonsky CJ, Hu Y, Huckins LM, Jeff J, Justice AE, Kocarnik JM, Lim U, Lin BM, Lu Y, Nelson SC, Park SL, Poisner H, Preuss MH, Richard MA, Schurmann C, Setiawan VW, Sockell A, Vahi K, Verbanck M, Vishnu A, Walker RW, Young KL, Zubair N, Acuña-Alonso V, Ambite JL, Barnes KC, Boerwinkle E, Bottinger EP, Bustamante CD, Caberto C, Canizales-Quinteros S, Conomos MP, Deelman E, Do R, Doheny K, Fernández-Rhodes L, Fornage M, Hailu B, Heiss G, Henn BM, Hindorff LA, Jackson RD, Laurie CA, Laurie CC, Li Y, Lin DY, Moreno-Estrada A, Nadkarni G, Norman PJ, Pooler LC, Reiner AP, Romm J, Sabatti C, Sandoval K, Sheng X, Stahl EA, Stram DO, Thornton TA, Wassel CL, Wilkens LR, Winkler CA, Yoneyama S, Buyske S, Haiman CA, Kooperberg C, Le Marchand L, Loos RJF, Matise TC, North KE, Peters U, Kenny EE, Carlson CS. Genetic analyses of diverse populations improves discovery for complex traits. Nature. 2019 Jun;570(7762):514–518. doi: 10.1038/s41586-019-1310-4. Epub 2019 Jun 19. PMID: 31217584; PMCID: PMC6785182.

46. Minster, R.L., et al. (2020). Evolutionary history of modern Samoans. Proceedings of the National Academy of Sciences, 117(17), 9458–9465. 10.1073/pnas.1913157117

47. Maskarinec G, Novotny R, Tasaki K. Dietary patterns are associated with body mass index in multiethnic women. J Nutr. 2000 Dec;130(12):3068–72. doi: 10.1093/jn/130.12.3068. PMID: 11110871.

48. Martínez-González MA, García-Arellano A, Toledo E, Salas-Salvadó J, Buil-Cosiales P, Corella D, Covas MI, Schröder H, Arós F, Gómez-Gracia E, Fiol M, Ruiz-Gutiérrez V, Lapetra J, Lamuela-Raventos RM, Serra-Majem L, Pintó X, Muñoz MA, Wärnberg J, Ros E, Estruch R; PREDIMED Study Investigators. A 14-item Mediterranean diet assessment tool and obesity indexes among high-risk subjects: the PREDIMED trial. PLoS One. 2012;7(8):e43134. doi: 10.1371/journal.pone.0043134. Epub 2012 Aug 14. PMID: 22905215; PMCID: PMC3419206.

49. Hurley KM, Oberlander SE, Merry BC, Wrobleski MM, Klassen AC, Black MM. The healthy eating index and youth healthy eating index are unique, nonredundant measures of diet quality among low-income, African American adolescents. J Nutr. 2009 Feb;139(2):359–64. doi: 10.3945/jn.108.097113. Epub 2008 Dec 11. PMID: 19074210; PMCID: PMC2646206.

50. Tande DL, Magel R, Strand BN. Healthy Eating Index and abdominal obesity. Public Health Nutr. 2010 Feb;13(2):208–14. doi: 10.1017/S1368980009990723. Epub 2009 Aug 4. PMID: 19650960.

51. Rudin C. (2019) Stop explaining black box machine learning models for high stakes decisions and use interpretable models instead. Nature Machine Intelligence 1:206–215. doi: 10.1038/s42256-019-0048-x

52. Lipton ZC. (2018) The mythos of model interpretability: In machine learning, the concept of interpretability is both important and slippery. Communications of the ACM 61(10):36–43. doi: 10.1145/3233231

53. Pan A, Sun Q, Bernstein AM, Schulze MB, Manson JE, Willett WC, Hu FB. Red meat consumption and risk of type 2 diabetes: 3 cohorts of US adults and an updated meta-analysis. Am J Clin Nutr. 2011 Oct;94(4):1088–96. doi: 10.3945/ajcn.111.018978. Epub 2011 Aug 10. PMID: 21831992; PMCID: PMC3173026.

54. Li C, Bishop TRP, Imamura F, Sharp SJ, Pearce M, Brage S, Ong KK, Ahsan H, Bes-Rastrollo M, Beulens JWJ, den Braver N, Byberg L, Canhada S, Chen Z, Chung HF, Cortés-Valencia A, Djousse L, Drouin-Chartier JP, Du H, Du S, Duncan BB, Gaziano JM, Gordon-Larsen P, Goto A, Haghighatdoost F, Härkänen T, Hashemian M, Hu FB, Ittermann T, Järvinen R, Kakkoura MG, Neelakantan N, Knekt P, Lajous M, Li Y, Magliano DJ, Malekzadeh R, Le Marchand L, Marques-Vidal P, Martinez-Gonzalez MA, Maskarinec G, Mishra GD, Mohammadifard N, O’Donoghue G, O’Gorman D, Popkin B, Poustchi H, Sarrafzadegan N, Sawada N, Schmidt MI, Shaw JE, Soedamah-Muthu S, Stern D, Tong L, van Dam RM, Völzke H, Willett WC, Wolk A, Yu C; EPIC-InterAct Consortium; Forouhi NG, Wareham NJ. Meat consumption and incident type 2 diabetes: an individual-participant federated meta-analysis of 1·97 million adults with 100 000 incident cases from 31 cohorts in 20 countries. Lancet Diabetes Endocrinol. 2024 Sep;12(9):619–630. doi: 10.1016/S2213-8587(24)00179-7. Erratum in: Lancet Diabetes Endocrinol. 2025 Feb;13(2):e2. doi: 10.1016/S2213-8587(25)00002-6. PMID: 39174161.

55. Malik VS, Popkin BM, Bray GA, Després JP, Willett WC, Hu FB. Sugar-sweetened beverages and risk of metabolic syndrome and type 2 diabetes: a meta-analysis. Diabetes Care. 2010 Nov;33(11):2477–83. doi: 10.2337/dc10-1079. Epub 2010 Aug 6. PMID: 20693348; PMCID: PMC2963518.

56. Jizhao Niu, Bai Li, Qing Zhang, Ge Chen, Angeliki Papadaki, Exploring the traditional Chinese diet and its association with health status—a systematic review, Nutrition Reviews, Volume 83, Issue 2, February 2025, Pages e237–e256, 10.1093/nutrit/nuae013

57. Boulangé, C.L., Neves, A.L., Chilloux, J. et al. Impact of the gut microbiota on inflammation, obesity, and metabolic disease. Genome Med 8, 42 (2016). 10.1186/s13073-016-0303-2

58. Jin Q, Black A, Kales SN, Vattem D, Ruiz-Canela M, Sotos-Prieto M. Metabolomics and Microbiomes as Potential Tools to Evaluate the Effects of the Mediterranean Diet. Nutrients. 2019 Jan 21;11(1):207. doi: 10.3390/nu11010207. PMID: 30669673; PMCID: PMC6356665.

59. Li D, Li Y, Yang S, Lu J, Jin X, Wu M. Diet-gut microbiota-epigenetics in metabolic diseases: From mechanisms to therapeutics. Biomed Pharmacother. 2022 Sep;153:113290. doi: 10.1016/j.biopha.2022.113290. Epub 2022 Jun 17. PMID: 35724509.

60. Putti R, Sica R, Migliaccio V, Lionetti L. Diet impact on mitochondrial bioenergetics and dynamics. Front Physiol. 2015 Apr 8;6:109. doi: 10.3389/fphys.2015.00109. PMID: 25904870; PMCID: PMC4389347.

61. Putti R, Sica R, Migliaccio V, Lionetti L. Diet impact on mitochondrial bioenergetics and dynamics. Front Physiol. 2015 Apr 8;6:109. doi: 10.3389/fphys.2015.00109. PMID: 25904870; PMCID: PMC4389347.

62. Niforou A, Konstantinidou V, Naska A. Genetic Variants Shaping Inter-individual Differences in Response to Dietary Intakes-A Narrative Review of the Case of Vitamins. Front Nutr. 2020 Dec 1;7:558598. doi: 10.3389/fnut.2020.558598. PMID: 33335908; PMCID: PMC7736113.

63. Sun H, Lin M, Russell EM, Minster RL, Chan TF, Dinh BL, Naseri T, Reupena MS, Lum-Jones A; Samoan Obesity, Lifestyle, and Genetic Adaptations (OLaGA) Study Group; Cheng I, Wilkens LR, Le Marchand L, Haiman CA, Chiang CWK. The impact of global and local Polynesian genetic ancestry on complex traits in Native Hawaiians. PLoS Genet. 2021 Feb 11;17(2):e1009273. doi: 10.1371/journal.pgen.1009273. PMID: 33571193; PMCID: PMC7877570.

64. Pacific islanders pay heavy price for abandoning traditional diet. Bull World Health Organ. 2010 Jul 1;88(7):484–5. doi: 10.2471/BLT.10.010710. PMID: 20616964; PMCID: PMC2897991.

65. Dela Cruz R, Wolfe E, Yonemori KM, Fialkowski MK, Wilkens LR, Coleman P, Lameko-Mua S, Johnson E, Gilmatam D, Sigrah C, Shomour M, Remengesau S, Alfred J, Acosta M, Ettienne R, Deenik J, Tanisha A, Salazar KA, Novotny R, Boushey CJ. Exploring Foods of the Pacific: Cultural Food Identity in the US Affiliated Pacific Region. Hawaii J Health Soc Welf. 2022 Sep;81(9):247–252. PMID: 36118154; PMCID: PMC9460762.

66. Lin M, Caberto C, Wan P, Li Y, Lum-Jones A, Tiirikainen M, Pooler L, Nakamura B, Sheng X, Porcel J, Lim U, Setiawan VW, Le Marchand L, Wilkens LR, Haiman CA, Cheng I, Chiang CWK. Population-specific reference panels are crucial for genetic analyses: an example of the CREBRF locus in Native Hawaiians. Hum Mol Genet. 2020 Aug 3;29(13):2275–2284. doi: 10.1093/hmg/ddaa083. PMID: 32491157; PMCID: PMC7399533.

67. Minster RL, Hawley NL, Su CT, Sun G, Kershaw EE, Cheng H, Buhule OD, Lin J, Reupena MS, Viali S, Tuitele J, Naseri T, Urban Z, Deka R, Weeks DE, McGarvey ST. A thrifty variant in CREBRF strongly influences body mass index in Samoans. Nat Genet. 2016 Sep;48(9):1049–1054. doi: 10.1038/ng.3620. Epub 2016 Jul 25. PMID: 27455349; PMCID: PMC5069069.

68. Stefan N, Häring HU, Schulze MB. (2020) Metabolically Healthy Obesity. Endocrine Reviews 41(3):bnaa004. doi: 10.1210/endrev/bnaa004

69. Vujkovic M, Keaton JM, Lynch JA, et al. (2024) Genetic drivers of heterogeneity in type 2 diabetes pathophysiology. Nature 627:347–357. doi: 10.1038/s41586-024-07019-6

70. Emmett PM, Jones LR. Diet, growth, and obesity development throughout childhood in the Avon Longitudinal Study of Parents and Children. Nutr Rev. 2015 Oct;73 Suppl 3(Suppl 3):175-206. doi: 10.1093/nutrit/nuv054. PMID: 26395342; PMCID: PMC4586450.

71. Solsona, E.M., Johnson, L., Northstone, K. et al. Prospective association between an obesogenic dietary pattern in early adolescence and metabolomics derived and traditional cardiometabolic risk scores in adolescents and young adults from the ALSPAC cohort. Nutr Metab (Lond) 20, 41 (2023). 10.1186/s12986-023-00754-z

72. Ambrosini GL, Emmett PM, Northstone K, Jebb SA. Tracking a dietary pattern associated with increased adiposity in childhood and adolescence. Obesity (Silver Spring). 2014 Feb;22(2):458–65. doi: 10.1002/oby.20542. Epub 2013 Sep 17. PMID: 23804590; PMCID: PMC3846445.

58. Foraita, R., Witte, J., Börnhorst, C. et al. A longitudinal causal graph analysis investigating modifiable risk factors and obesity in a European cohort of children and adolescents. Sci Rep 14, 6822 (2024). 10.1038/s41598-024-56721-y

73. Laska MN, Murray DM, Lytle LA, Harnack LJ. Longitudinal associations between key dietary behaviors and weight gain over time: transitions through the adolescent years. Obesity (Silver Spring). 2012 Jan;20(1):118–25. doi: 10.1038/oby.2011.179. Epub 2011 Jun 23. PMID: 21701567; PMCID: PMC3402912.

